# From tutor to future educator: Investigating the role of peer-peer tutoring in shaping careers in medical education

**DOI:** 10.1101/2023.12.05.23297167

**Authors:** Lauren Stokes, Harinder Singh

## Abstract

In the effort to promote academic excellence and provide authentic teaching experiences and training for medical students, the University of California, Irvine, School of Medicine (UCISOM) built a novel peer tutoring program, Collaborative Learning Communities with Medical Students as Teachers (CLC-MSAT). While the role of peer-assisted learning in student success on academic courses is well established, we wanted to assess the impact of our peer-assisted learning program on tutor’s career interest in medical education. Through a mixed-methods analysis of our peer tutors’ experiences, we found >80% were overall satisfied with their positions; >85% learned new skills; >88% felt they were strong teachers; >88% felt they now had a stronger grasp of the medical curriculum. Our findings suggest that the CLC-MSAT program has a positive impact in exposing medical students to teaching experience and training, which may help prepare the next generation of academic clinicians.

## Introduction

Peer assisted learning (PAL) refers to an education method in which a more senior-standing student provides instruction for their peers^1,2^. As an increasingly utilized teaching modality in medical education, PAL programs differ in size, duration, courses served, and resource availability, making the outcomes difficult to compare across institutions^1^. Strengths of PAL programs may include fostering a safe learning environment and providing mentorship opportunities, which are especially crucial for academically at-risk students^1^. For example, a medical education institution found that students who were most at risk of failing an undergraduate medical course had significant improvements in their final course grade after 3-4 months and 8-9 months of tutoring^3^.

Multiple studies have indicated that PAL programs have positive impacts for both tutors and tutees. For example, Wong et al. (2007) found that tutors had significantly higher USMLE scores on Step 1 and Step 2 CK, as well as GPAs^4^. These findings were consistent with outcomes for an international medical school, which demonstrated a positive relationship between the number of courses a tutor led and their overall medical school GPA^5^. Additionally, some universities have found that for objective measures such as examination scores and course grades, students who participated in peer tutoring performed better, on average, than their non-participant peers^6,7,8^. For more subjective measures, such as collaboration and leadership skills, peer tutoring has been found to also have a positive influence on tutors^9,10,11^.

Within the undergraduate medical education field, one study found the students who were most likely to participate in their services included students who identified as female, underrepresented, and/or disadvantaged, highlighting the importance for institutions to provide inclusive and high quality peer tutoring resources^12^. Strengths of PAL programs also include the prospect that peers are able to more accurately determine knowledge gaps compared to teacher-led learning, as well as creating a horizontal hierarchy in the teaching environment^13,14,15^. PAL programs are deemed as a bidirectional and reciprocal process in which the tutor naturally discovers their own level of content understanding based on their ability to teach the content to another person^16,17^. This type of educational strategy is meant to supplement teacher-led learning as an additional resource to serve the wide learning levels of a student cohort^18^.

The peer tutoring program entitled, Collaborative Learning Communities with Medical Students as Teachers (CLC-MSAT) program at UCISOM launched in 2019 and has successfully evolved to serve first through third year medical students^19^. As the team grew from a staff of 18 to 73 peer tutors in a four year period, our attention has also turned into cultivating future academic-oriented clinicians interested in exploring medical education as a career. Specifically, our team is invested in providing timely, relevant, and ongoing professional development in evidence-based teaching pedagogy, diversity, equity, and inclusion (DEI) training, as well as mental health advocacy. For example, MSATs are required to participate in a suicide prevention certification training, referred to as Question, Persuade, Refer (QPR), which helps MSATs determine the signs of students they may work with combatting mental health challenges.

The focus of this paper is to provide an understanding of our MSATs’ experiences serving within the CLC-MSAT program, including their overall satisfaction, recommended areas for improvement, and the potential impact their teaching roles have on future career aspirations.

## Methods

### Ethics Statement

The study was approved by University of California, Office of Research, Human Research Protections, Institutional Review Board (IRB), Kauli Research Protocols (KRP), Protocol *#3183* and was deemed “Exempt” upon review (IRB Approval form attached in supplementary data)

### Instrument

Our research team developed an inventory to help understand the impact that serving as an MSAT has had on our students, including how the experience has potentially impacted their career aspirations. Students who served as MSATs during the 2022-2023 academic year were invited to participate in the survey in May 2023. The inventory explored if the MSAT had prior teaching experience or formal teaching training before serving within the CLC program, satisfaction with their position, collaboration experiences with other MSATs and students, and how this role potentially had any impact on their career trajectory.

In addition to demographic information and previous teaching experience, the survey had a total of nine likert scale items and three open-ended items to assess satisfaction, career trajectory, and areas for continued growth of the program. MSATs were invited to complete the survey by the staff director of the program at the end of the third iteration of the program in 2023. All participation was voluntary and anonymous.

### Previous Teaching Experience

In the effort to gauge previous teaching experience or formal training, MSATs were asked if their experience or training took place at the K-12, undergraduate, or graduate level, as well as “no previous experience or training.” A total of nine statements assessed satisfaction and career trajectory items that were based on a 1 - 5 Likert scale with a 1 indicating a strongly disagree response to the statement, whereas a 5 indicated a strong agreement. Single-question items included: *Overall, I am satisfied with my position as an MSAT, I would recommend for other students to serve as an MSAT,* and *I have learned new skills in this position*.

To reflect on how the role has influenced their perceptions as a teacher, MSATs were asked: *after serving as an MSAT, I feel I am a stronger teacher,* as well as*, I have a stronger grasp of the medical school curriculum*. To address the notion of collaboration and effective teaching, two items included:

a. *I work well with my colleagues in this program*
b. *I work well with my students in this program*

To measure how the CLC program has potentially influenced the MSATs’ career trajectories, they were asked to rate:

a. *Before I served as an MSAT, I envisioned a career in academic medicine*
b. *Now that I have served as an MSAT, I envision a career in academic medicine*.

### Open-Ended Items

In addition to the Likert scale items, three open-ended questions asked the following:

a. *What are some of the aspects you enjoyed most about serving as an MSAT?*
b. *Has serving as an MSAT had any influence on your career aspirations?*

Respondents were also provided with the opportunity to disclose areas for improvement in an item which posed:

a. *How can the CLC-MSAT program improve to better support MSATs?*
b. *Did you encounter any specific challenges as an MSAT?*

Finally, the third open-ended item requested for *any additional comments, concerns, or suggestions the MSAT wanted to share with the research team*.

### Demographic Items

MSATs were invited to disclose non-identifying demographic information, such as their current year and program in medical school, age, gender, and first or second college generation status. All demographic questions had the option of “*prefer not to say*” to ensure respondents did not feel any identifying information was being solicited.

## Results

### Demographics of Respondents

A total of 28 MSATs completed the MSAT Feedback Survey in May 2023 out of a team of 43 (response rate= 65%). The academic years the respondents indicated serving as an MSAT were as follows: 2020-2021, 25% (*n*= 7), 2021-2022, 32% (*n*= 9), and 2022-2023, 96%, (*n*= 27) (Figure 1).

**Figure 1.**
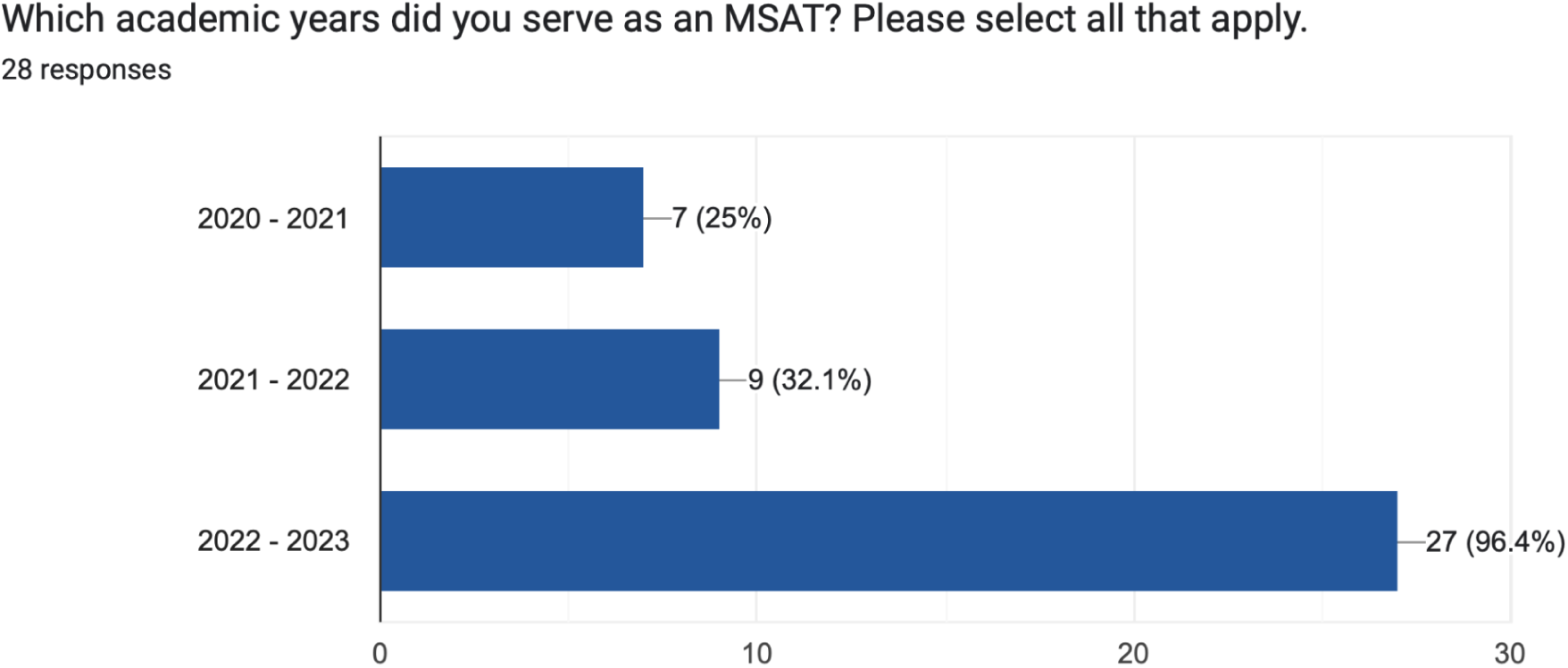
Academic years during which students served as MSATs

The breakdown of class standings of MSATs were: 43%= MS4, 39%= MS3, 14%= “Other” (Leave of absence or different educational track), and 3%= MS2 (Figure 2).

**Figure 2.**
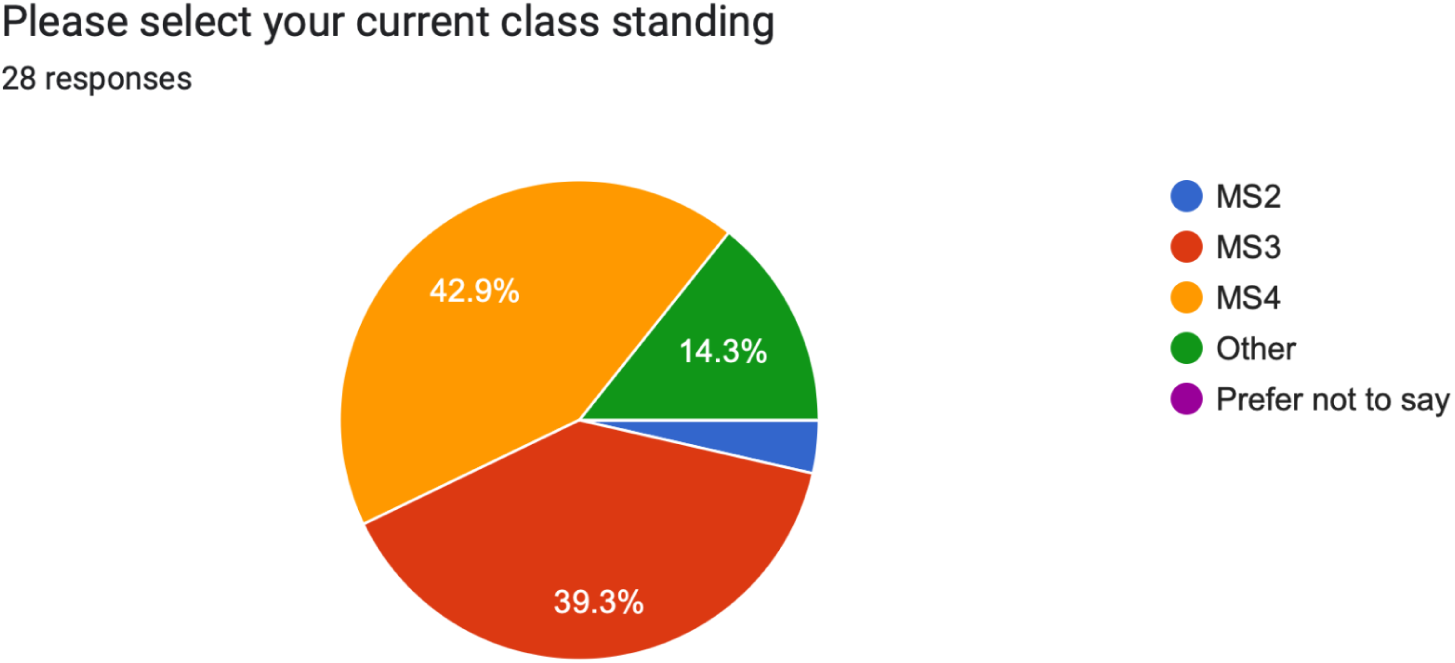
Breakdown of stage in medical education

Most participants were MD-exclusive students (79%) compared to 11% of MD/PhD, 7% MD/MBA, and 4% MD/MPH (Figure 3).

**Figure 3.**
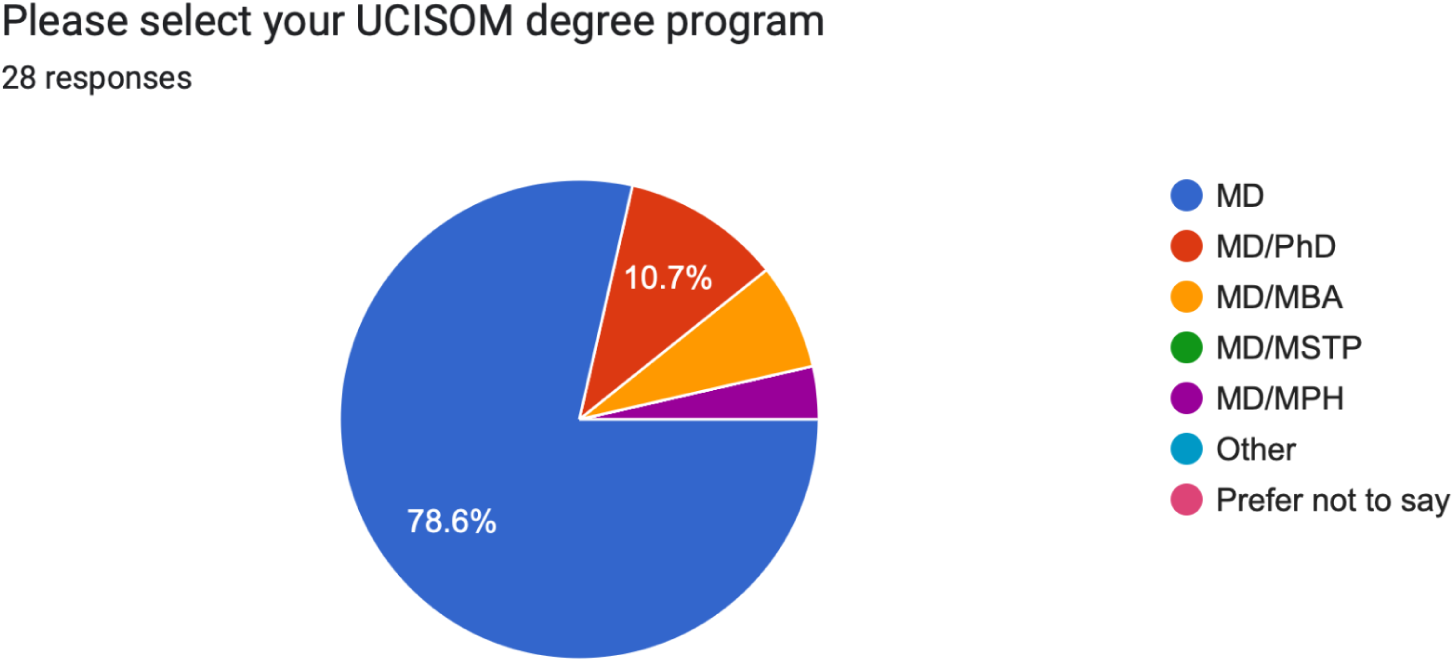
Degree program enrollment of MSATs

Additionally, 18% of respondents were students in one of the mission-based programs at UCISOM, which include LEAD PRIME-LC and PRIME LEAD-ABC compared to 79% of students not in a mission-based program. PRIME-LC refers to the Program in Medical Education for the Latino Community, which trains future physicians to meet the needs of under-resourced Latino communities. PRIME LEAD-ABC refers to the Program in Medical Education Leadership Education to Advance Diversity African, Black, and Caribbean. PRIME LEAD-ABC produces physician leaders who address the diverse health needs of Black communities. One student indicated “prefer not to say” regarding their mission-based program status or not (Figure 4).

**Figure 4.**
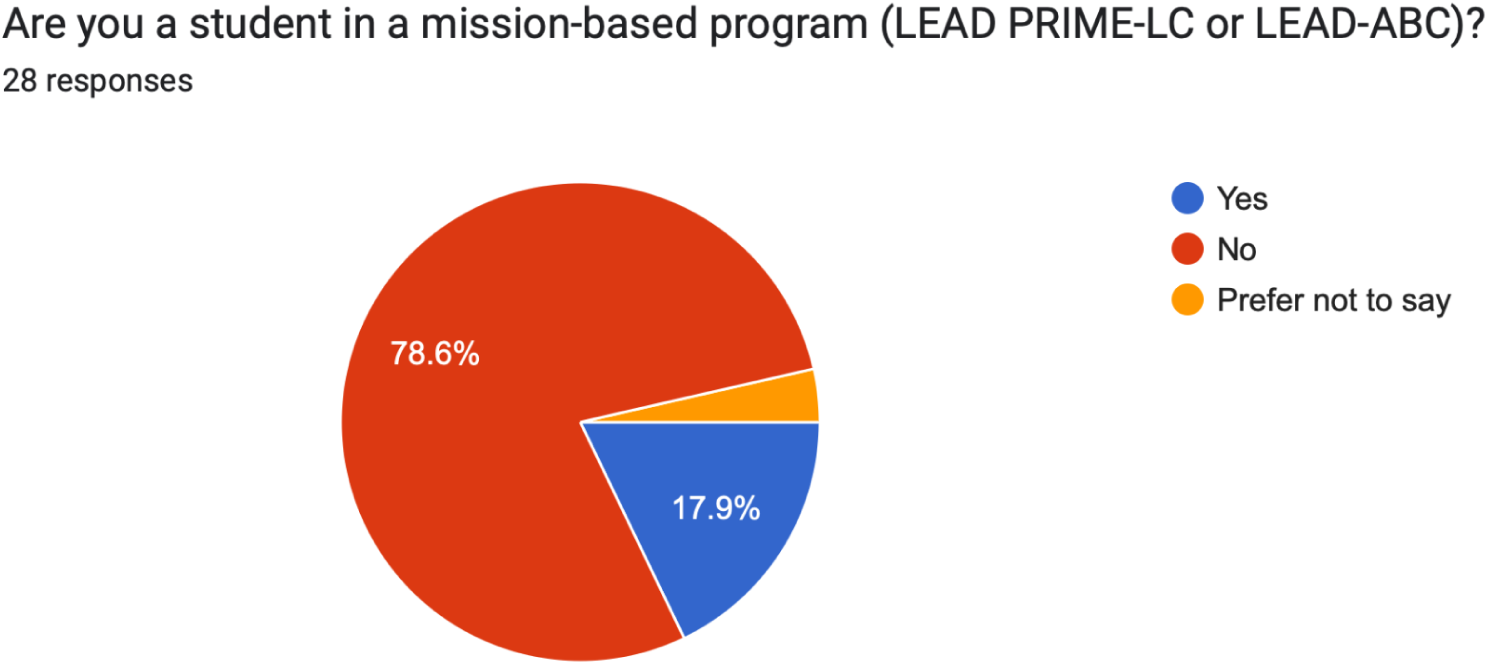
MSAT affiliation with mission-based programs at UCI-SOM.

Since the inception of CLC in 2020, several upper class-standing students have served in multiple CLC-level programs over the past three years. Most participants (71%) served in the MS1 program, compared to 39% in the MS2 program, 39% in the MS3 program, and 4% who preferred not to disclose which program they had ever served as an MSAT (Figure 5).

**Figure 5.**
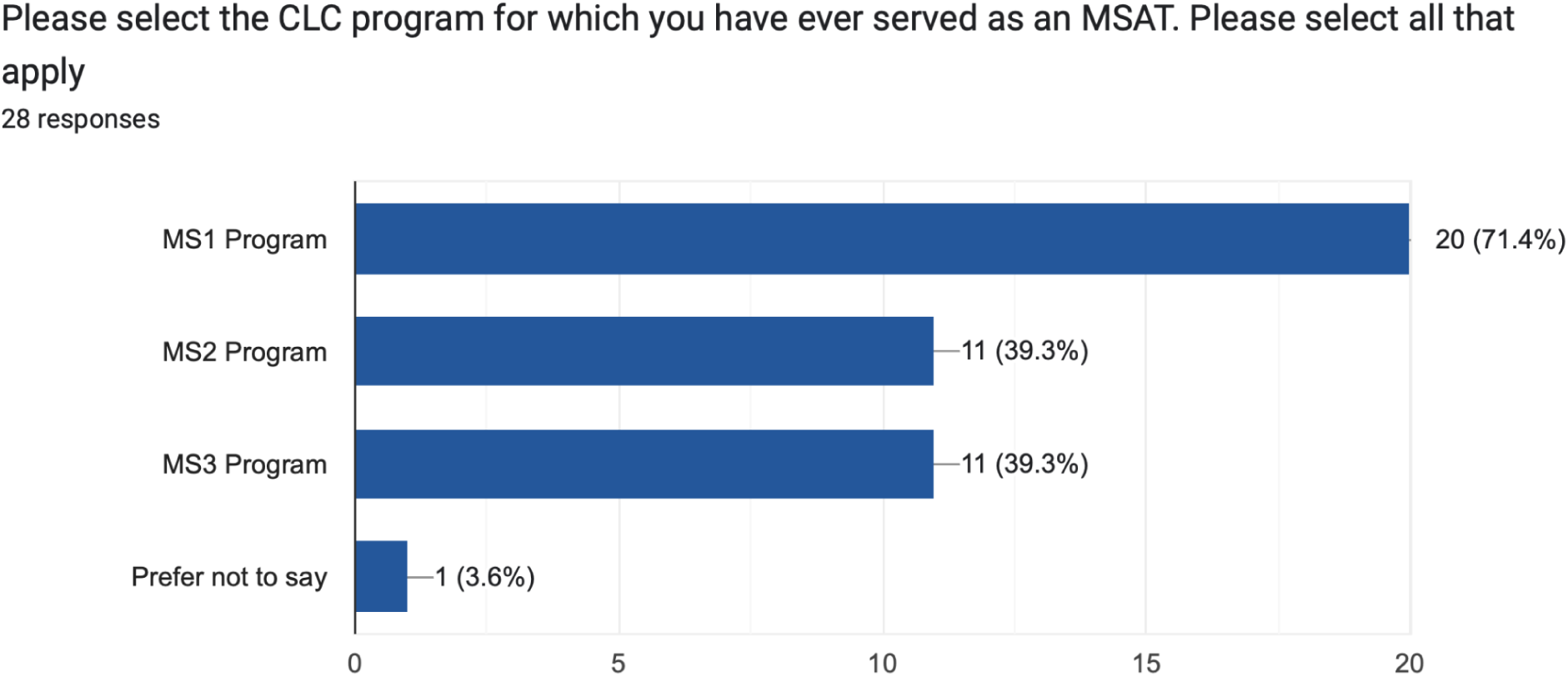
MSATs serving in CLC programs during their medical education years.

While 68% of respondents served as an MSAT-only (*n*= 19), 25% held a student director level role (*n*= 7), 25% held a specialist position (*n*= 7), 11% indicated an “Other” position (*n*= 3), and 4% preferred not to say (*n*= 1) (Figure 6).

**Figure 6.**
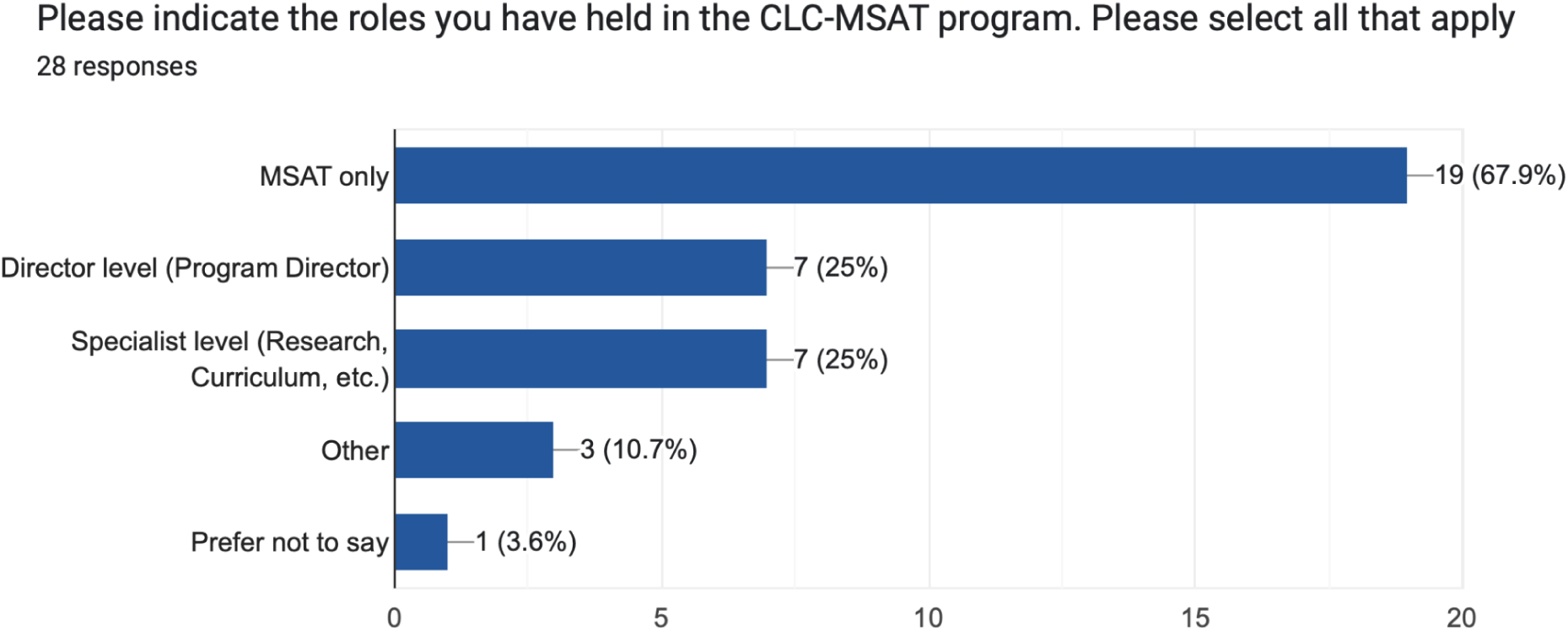
Leadership roles held by MSATs in the program.

Based on age range, 64% of MSATs indicated they were between 26-29 years old, 29% were between 22-25 years old, and 7% were between 30-33 years old (Figure 7).

**Figure 7.**
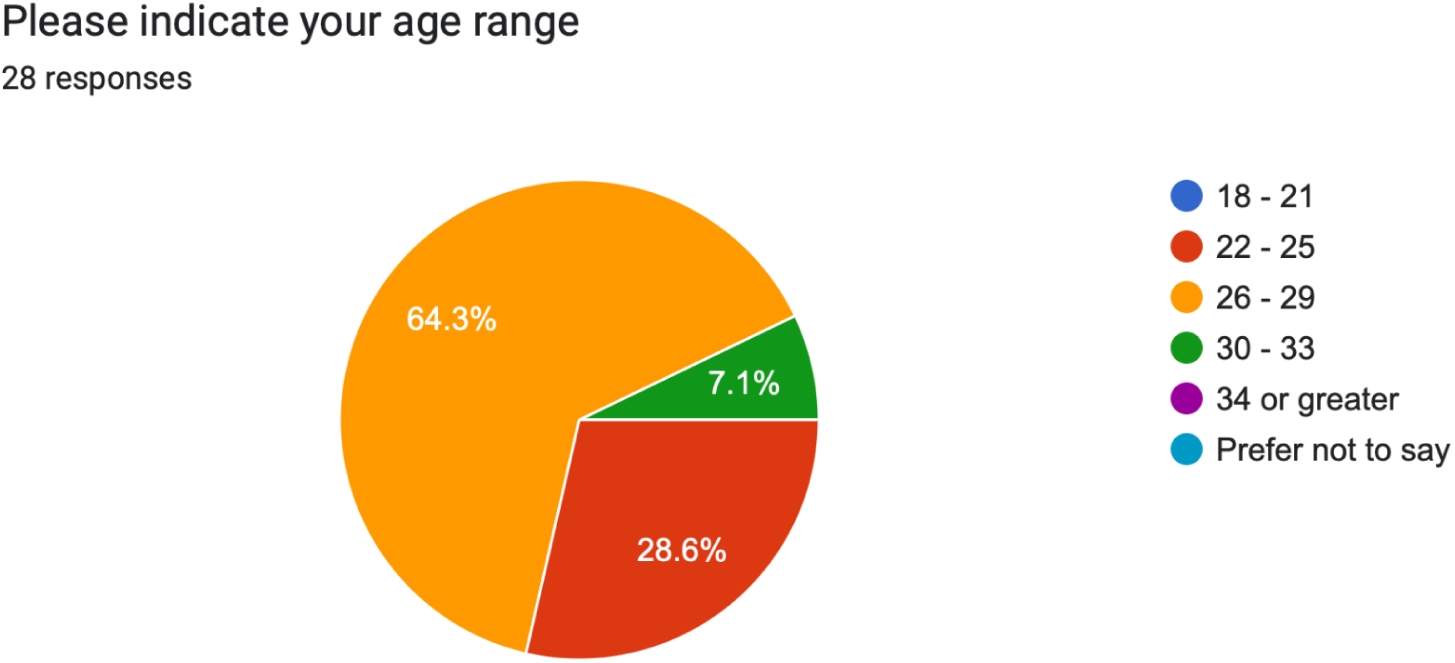
Age groups of CLC-MSATs

Exactly 50% of the participants identified as male (*n*= 14) compared to 50% who identified as female (*n*= 14) (Figure 8).

**Figure 8.**
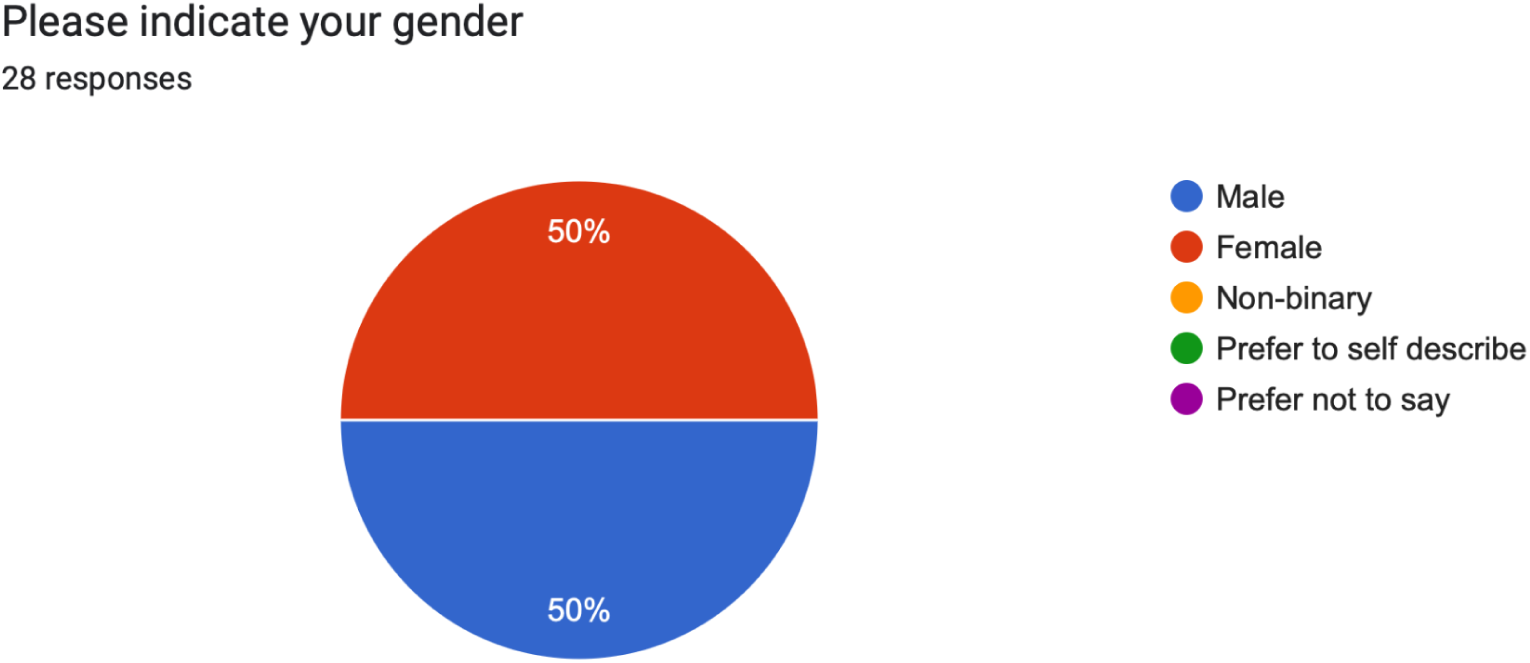
Gender of MSATs.

Based on college-generation status, 96% of MSATs were a second generation college student compared to 4% who were the first in their immediate family to attend college (Figure 9).

**Figure 9.**
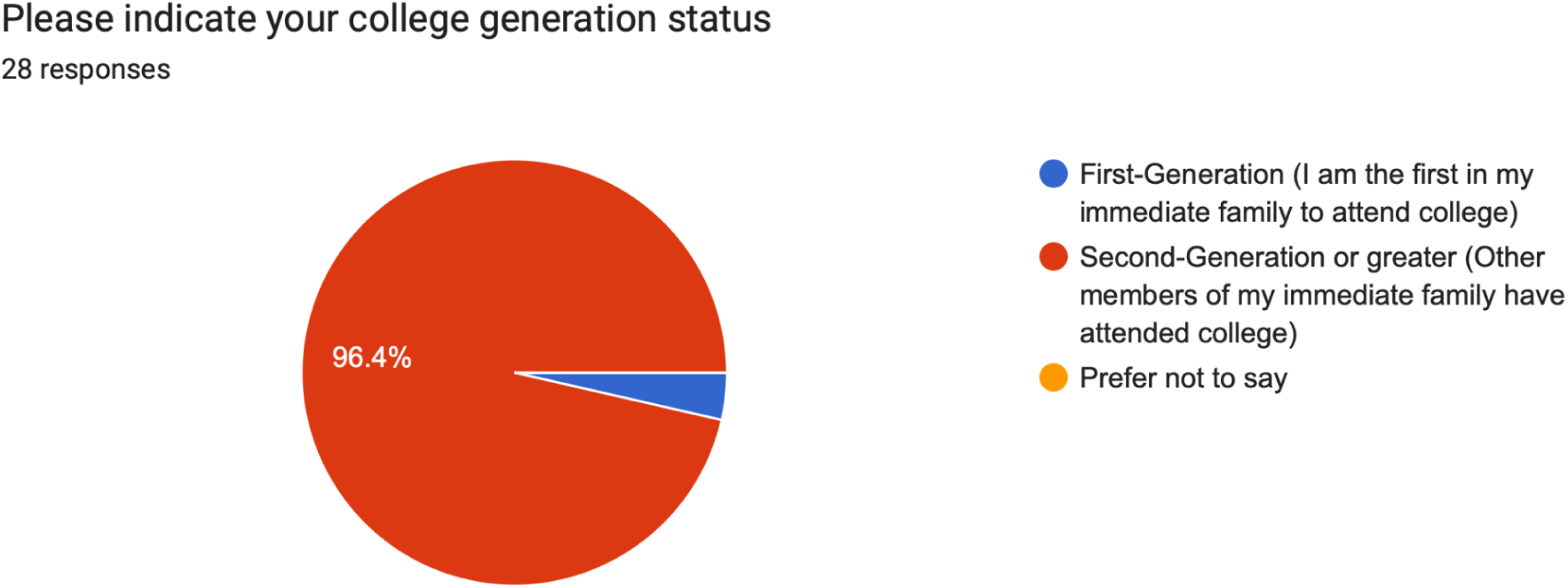
College generation status.

### Professional Preparation

Based on prior teaching experience, 74% of MSATs had served in a teaching capacity at the undergraduate level (*n*= 20), 59% at the K-12 level (*n*= 16), 15% in graduate school (*n*= 4), and 11% of the MSATs had no prior teaching experience before the CLC-MSAT program (*n*= 3) (Figure 10).

**Figure 10.**
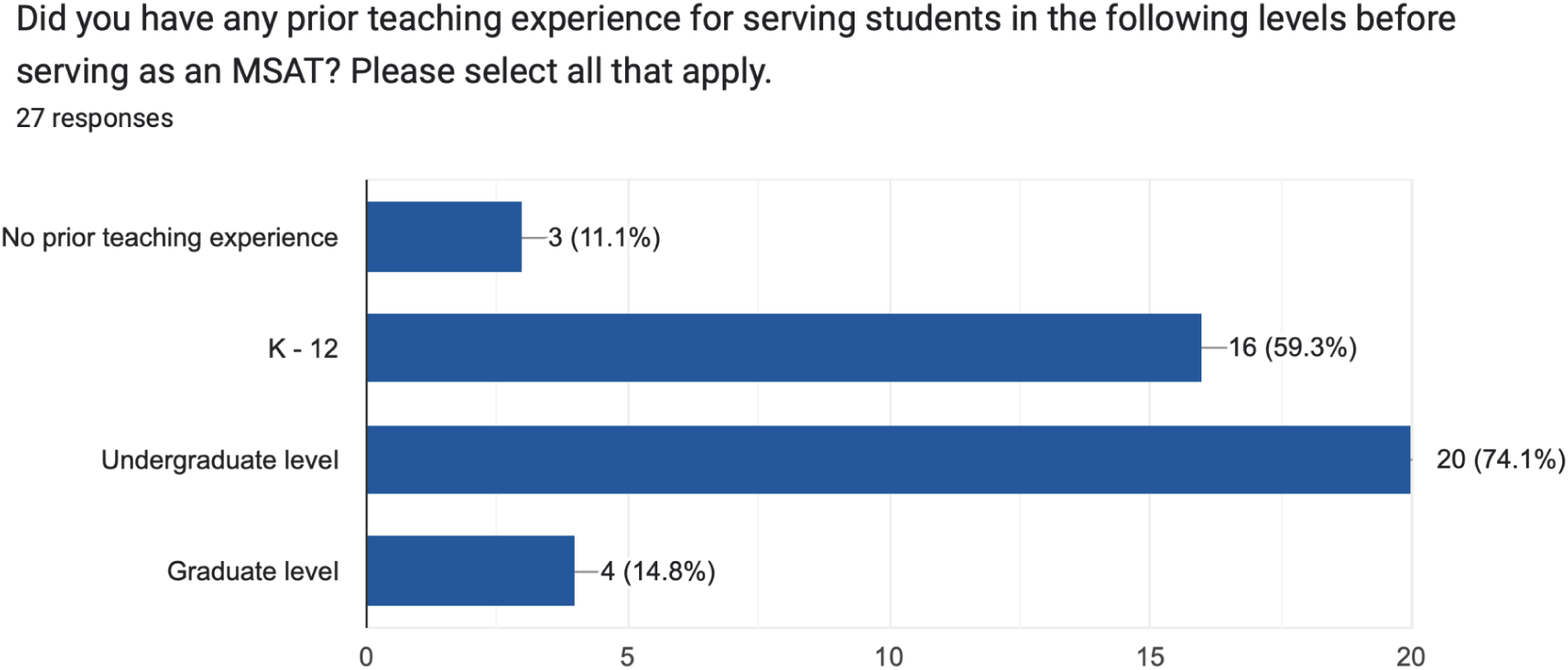
MSATs with prior teaching experience at different levels.

Nearly half of the MSATs had no formal teaching training (48%) compared to 19% to teach at the K-12 level, 41% for undergraduate students, and 11% for graduate students (Figure 11).

**Figure 11.**
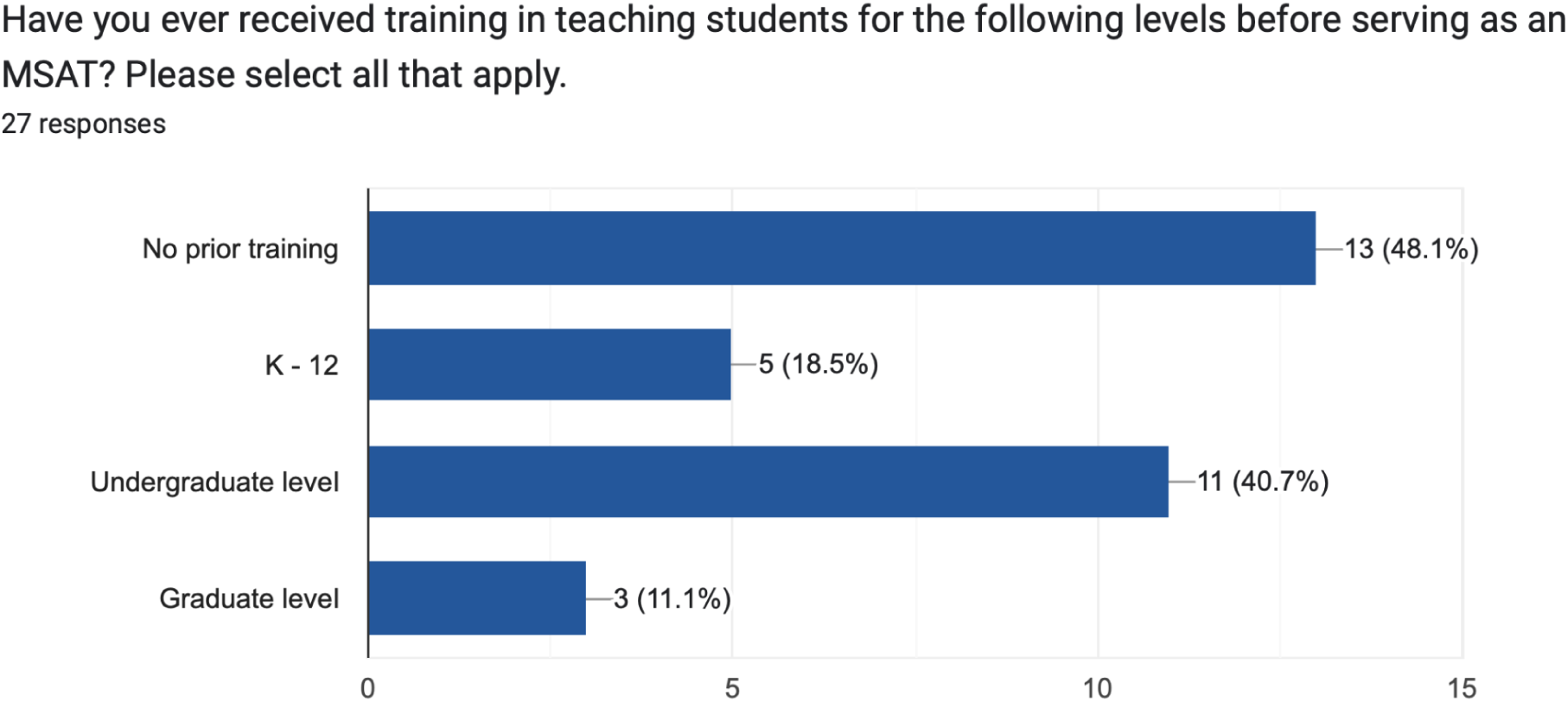
MSATs with prior training in teaching at different levels.

### MSAT Satisfaction

MSAT satisfaction was measured on a 1-4 Likert scale with a 1 indicating a strong disagreement and a 4 indicating a strong agreement. When asked to rate their overall satisfaction as an MSAT, 68% (*n*= 19) indicated they strongly agreed they were satisfied compared to 32% (*n*= 9) who agreed to the same statement. No MSATs indicated a disagreement or strong disagreement regarding their satisfaction (Figure 12).

**Figure 12.**
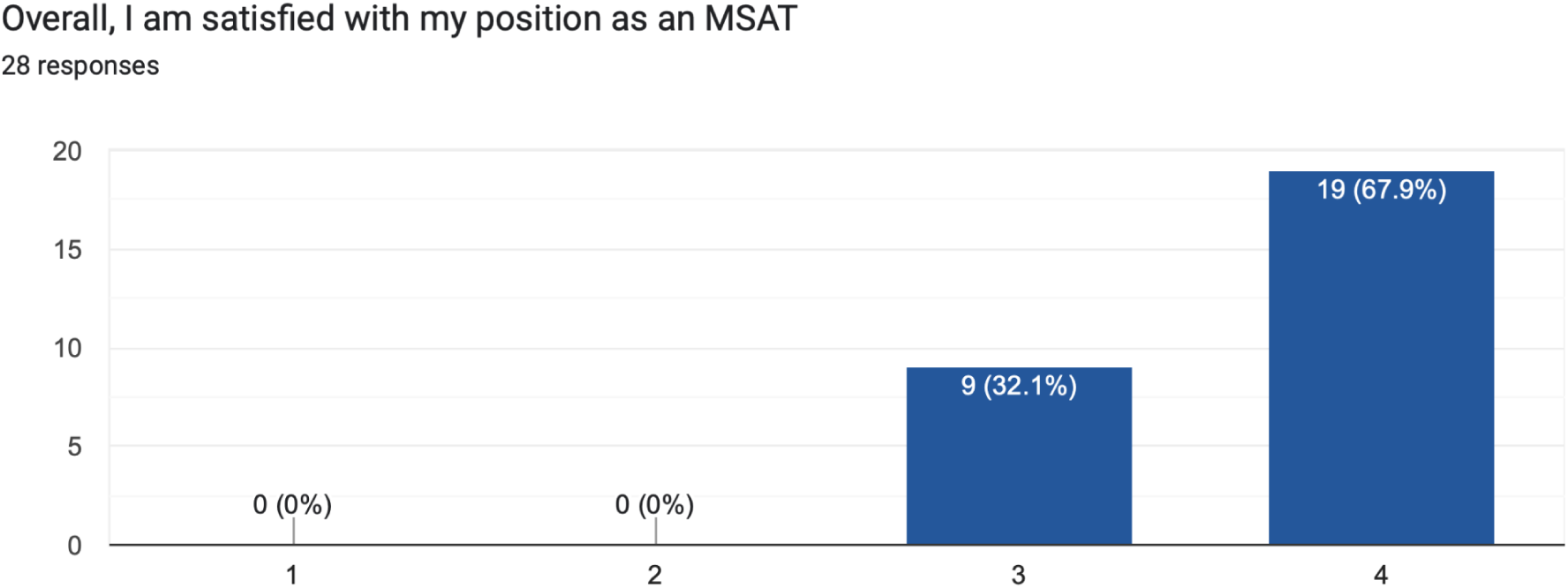
Overall satisfaction of MSATs with their position

A total of 76% of MSATs strongly agreed (71%) or agreed (25%) they would recommend for other students to serve as an MSAT compared to one student (4%) who indicated they disagreed with this statement (Figure 13).

**Figure 13.**
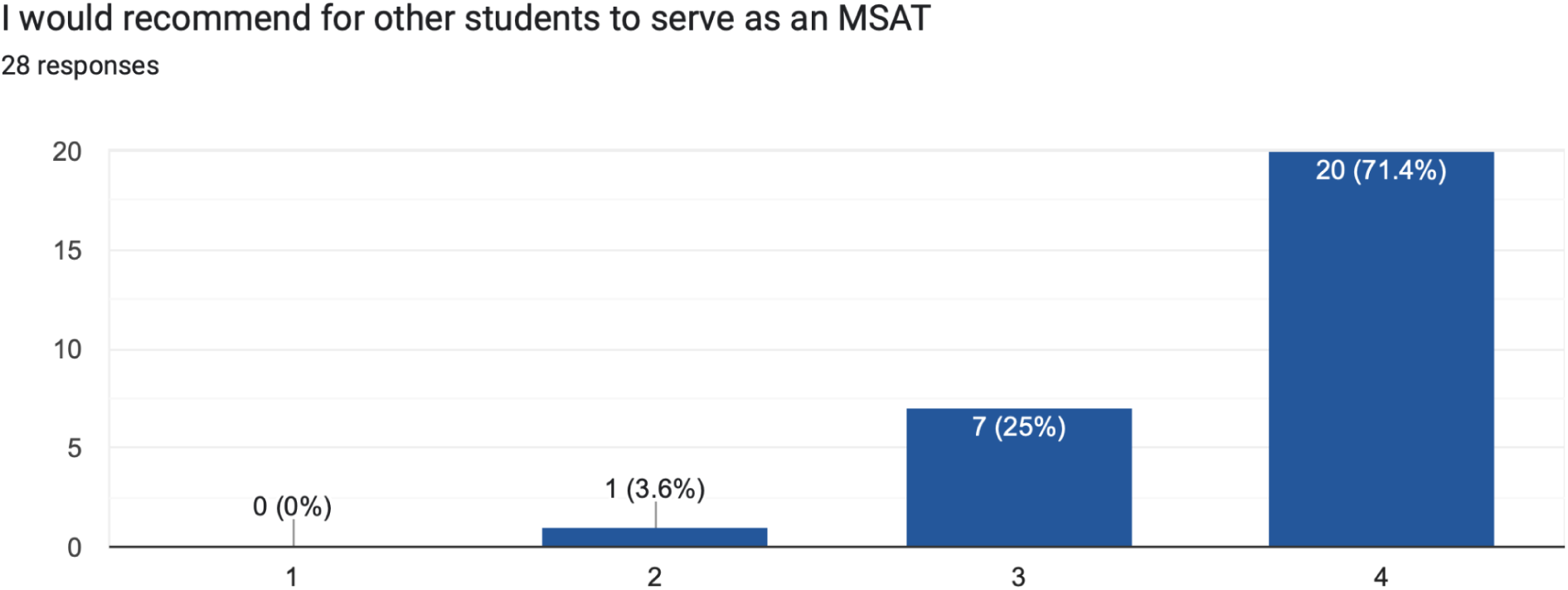
Recommendation for other students to enroll into the program.

### Personal Development

To measure personal and professional development, 86% of respondents strongly agreed (50%) or agreed (36%) they learned new skills in the position compared to 14% who disagreed (Figure 14).

**Figure 14.**
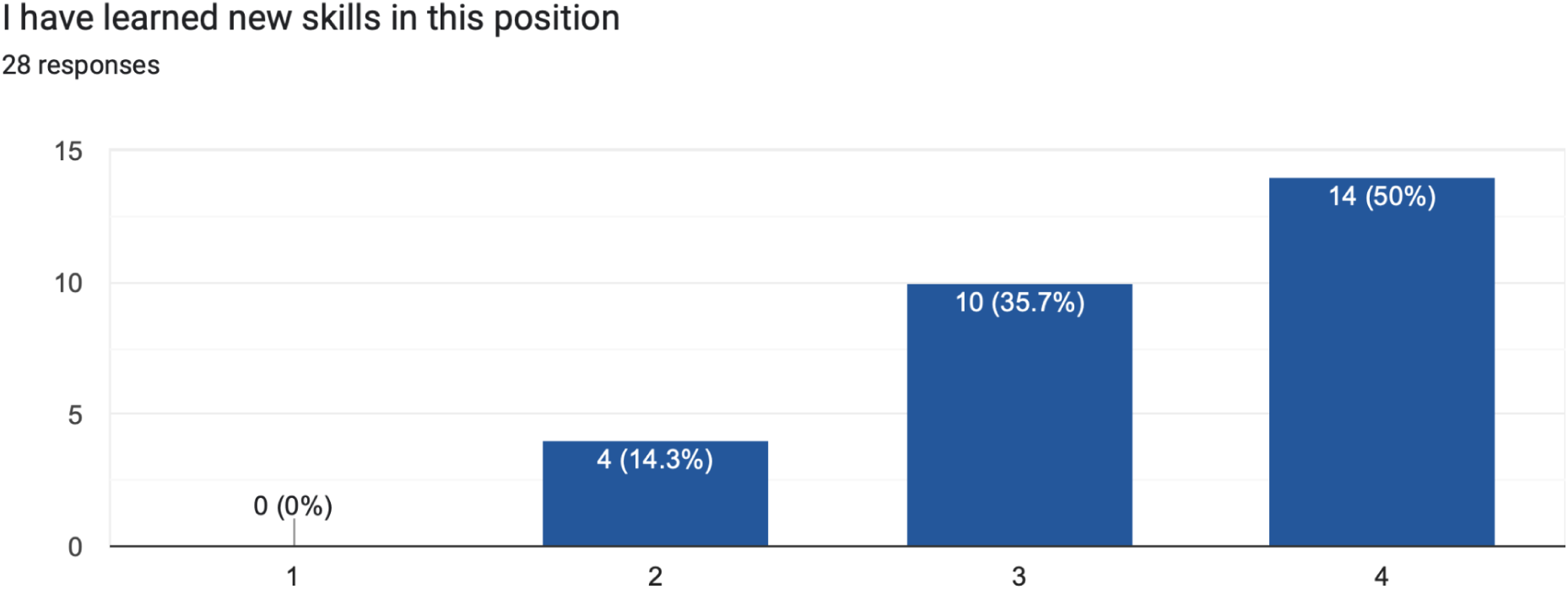
Development of professional and personal skills development.

Most MSATs (89%) strongly agreed (61%) or agreed (29%) they were a stronger teacher after serving as an MSAT compared to 11% who disagreed (Figure 15).

**Figure 15.**
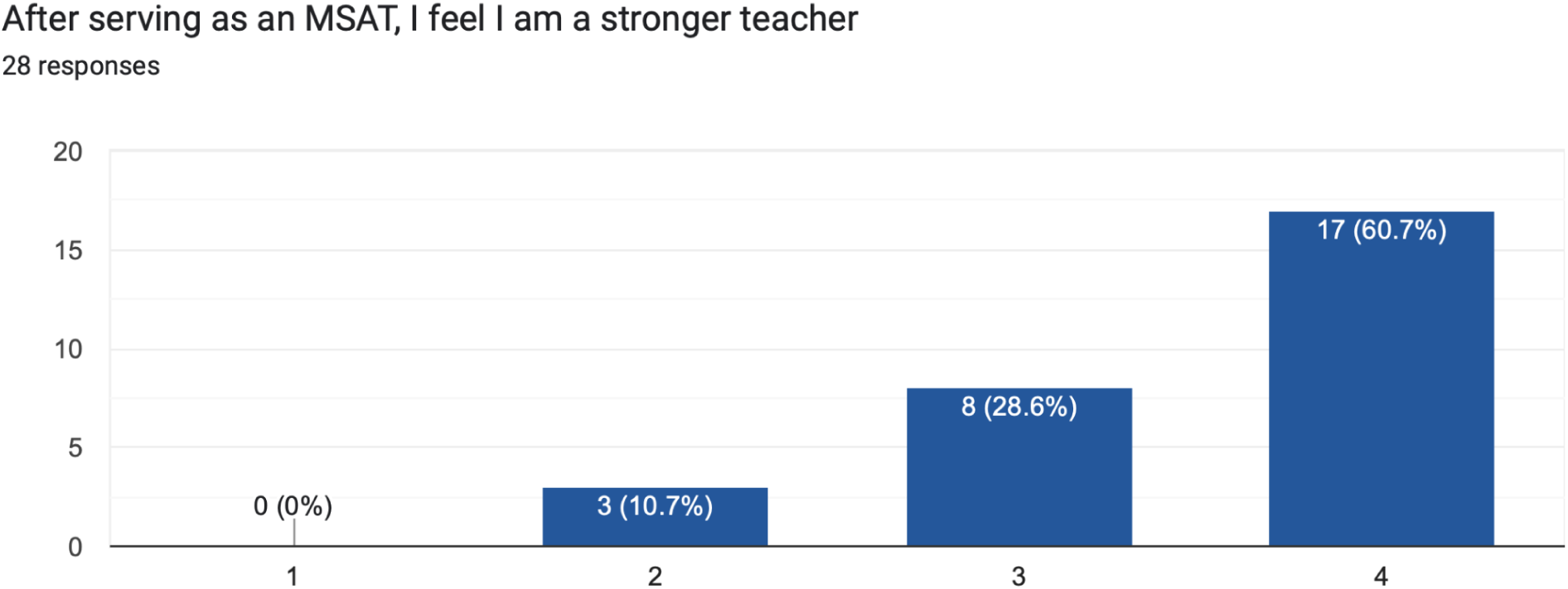
Perceptions as a stronger teacher after participating in the program.

**Figure 16.**
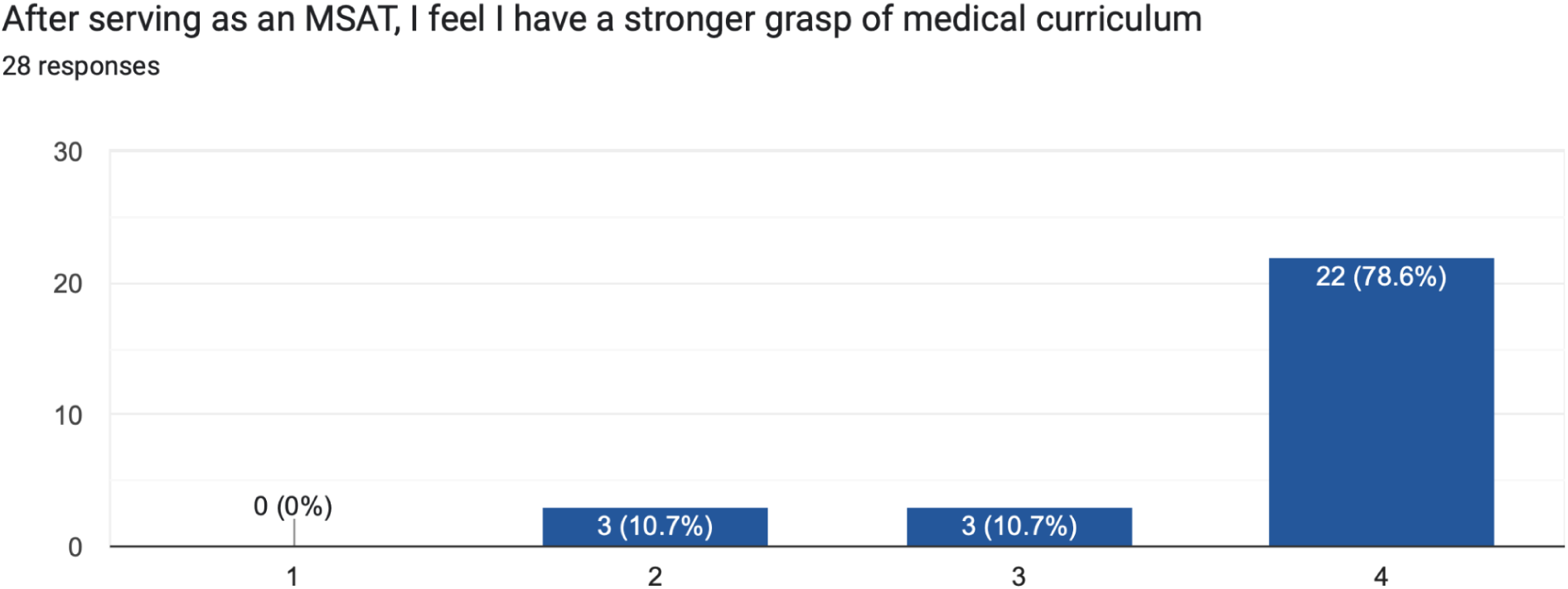
Grasp level of medical curriculum after participating in the program.

An item to measure if MSATs gained a stronger grasp of the medical curriculum as a result of serving as an MSAT, 89% strongly agreed or agreed to the statement compared to 11% who disagreed.

### Collaboration Effectiveness

MSATs were asked to gauge their effectiveness in collaborating with their colleagues and students in the program. Most MSATs (86%) strongly agreed (78%) or agreed (19%) they worked well with their colleagues compared to 4% who disagreed (Figure 17).

**Figure 17.**
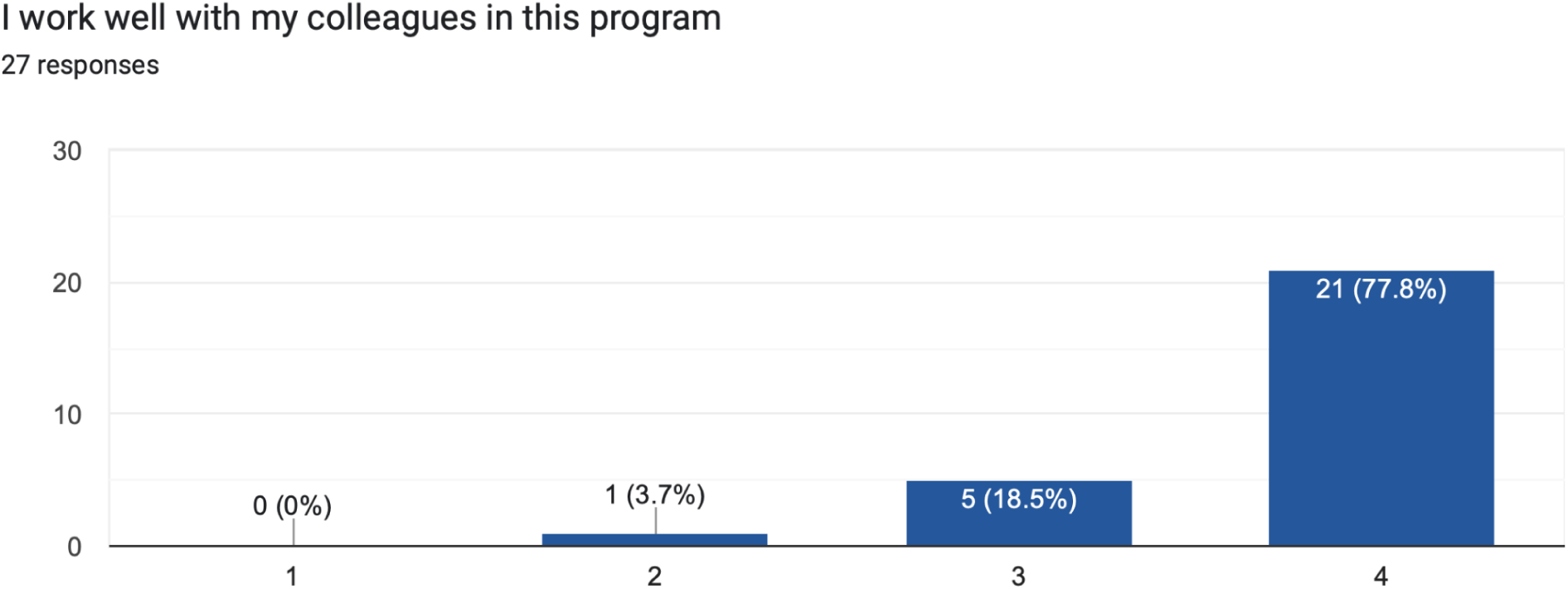
Collaboration spirit of students in the program.

In comparison, all MSATs (100%) strongly agreed (70%) or agreed (30%) they worked well with their students (Figure 18).

**Figure 18.**
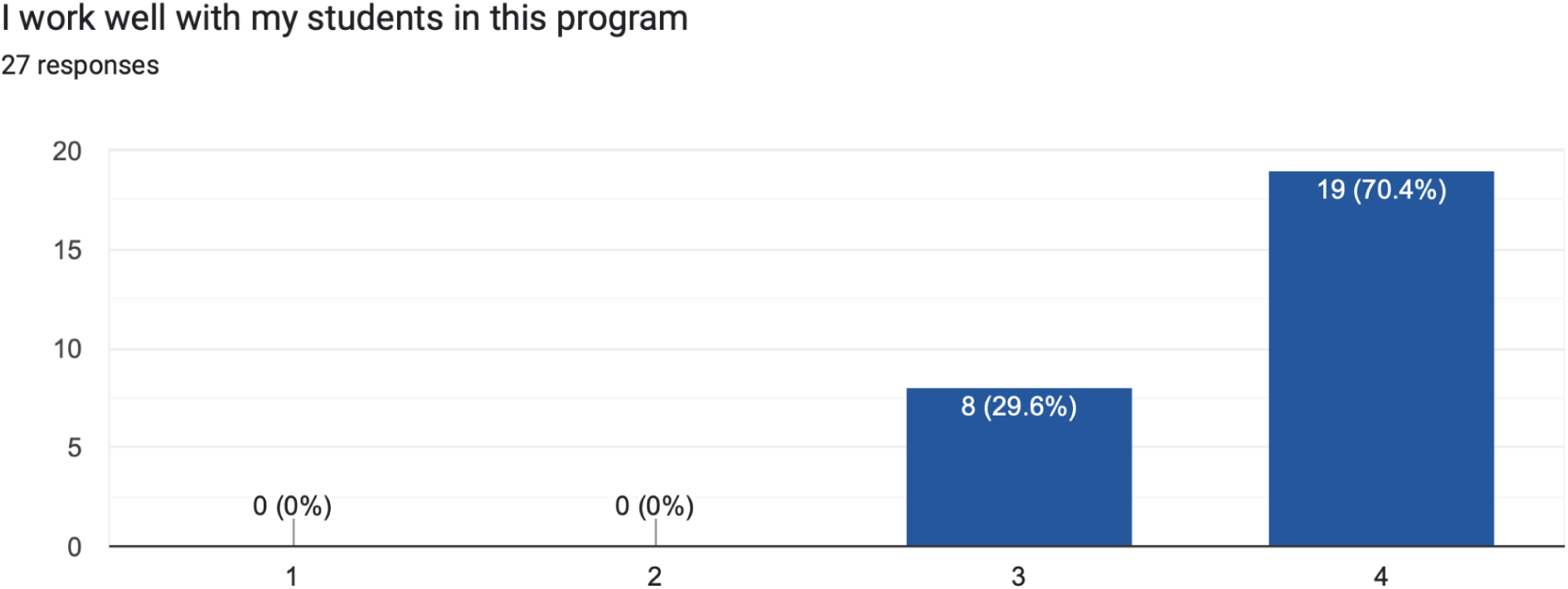
Ease of working well with others in the program.

### Career Trajectory

To measure the effect serving as an MSAT has had on their career trajectory, MSATs were asked to rate how they envisioned a career in academic medicine before serving in the CLC program. While most MSATS (89%) strongly agreed (61%) or agreed (29%), about 11% of the team disagreed (7%) or strongly disagreed (4%) that they envisioned a career in academic medicine before serving as an MSAT (Figure 19).

**Figure 19.**
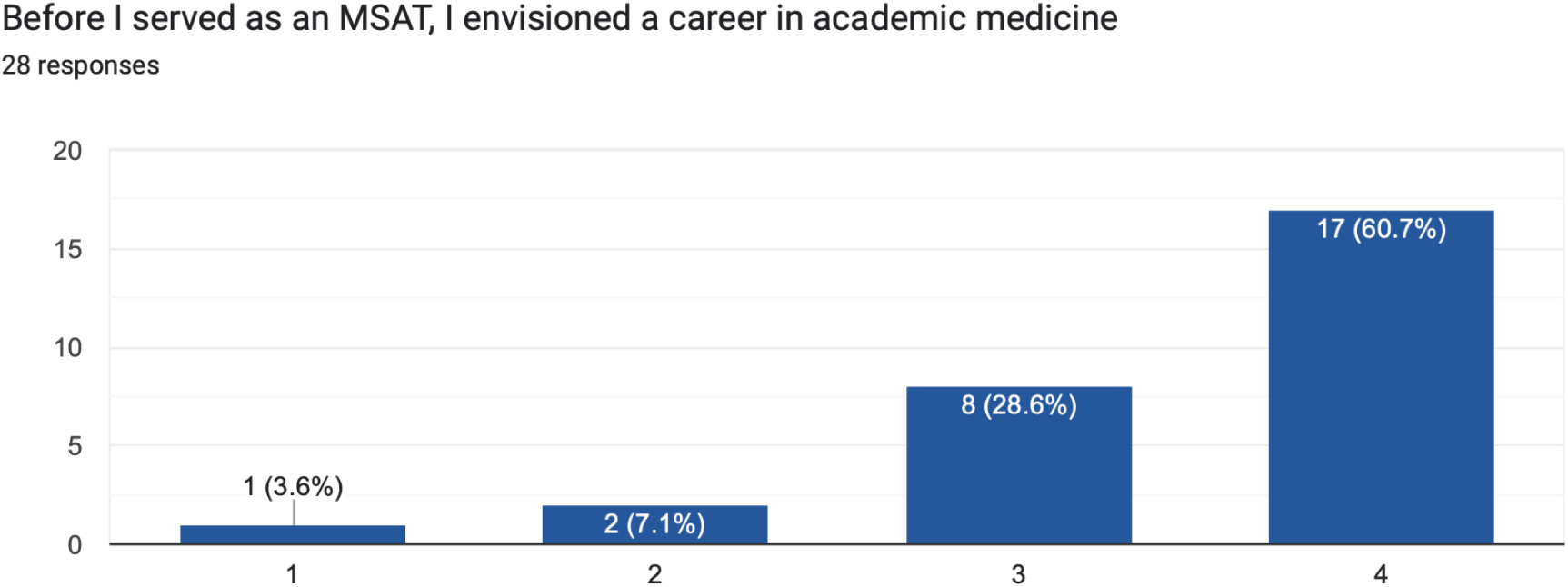
Interest in a career in academic medicine before starting the MSAT program

After serving as an MSAT, 96% of the team either strongly agreed (63%) or agreed (33%) they envisioned a career in academic medicine compared to 4% who disagreed. No respondents strongly disagreed with this statement (Figure 20).

**Figure 20.**
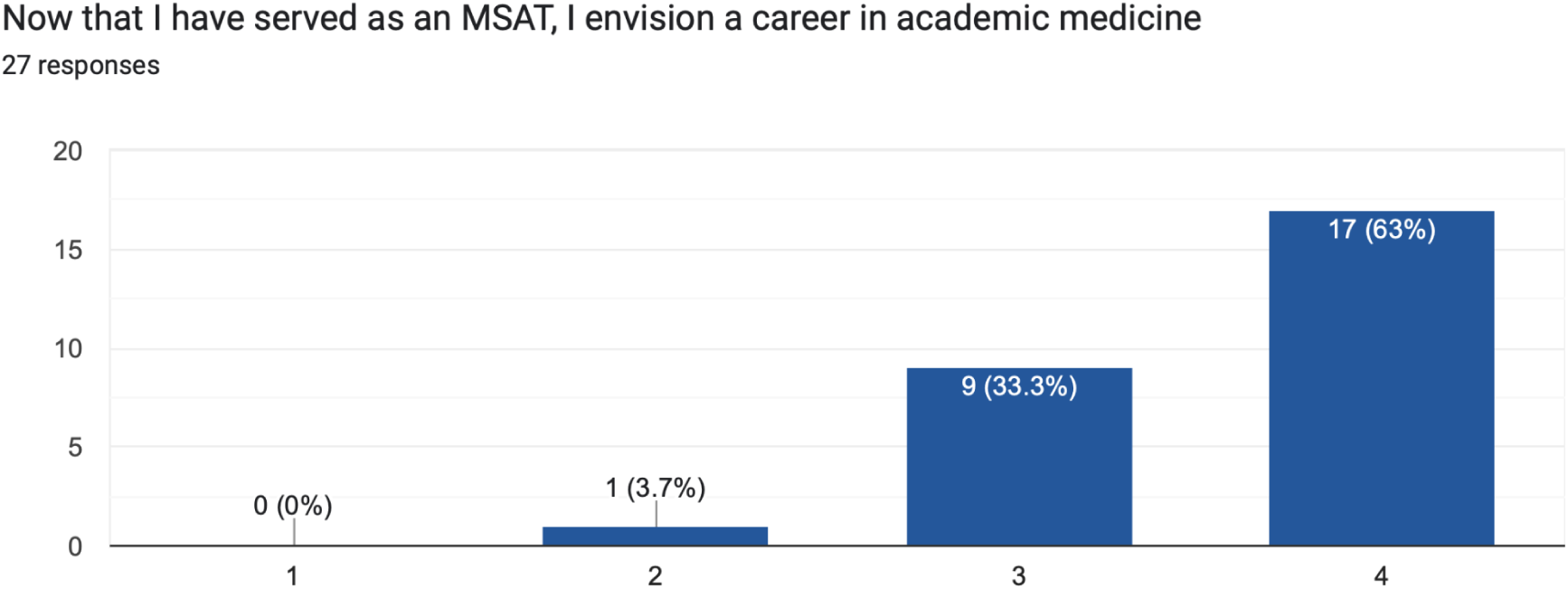
Interest in a career in academic medicine after serving in the MSAT program.

### Open-Ended Items

Three open-ended items invited MSATs to disclose their thoughts about the specific aspects they enjoyed while serving in CLC, as well as how this opportunity has potentially impacted their career trajectory. Recommendations for program improvement were solicited with an additional platform to disclose any remaining comments and concerns. General themes are presented below.

The first open-ended item invited MSATs to share the aspects they enjoyed most about serving in the program (Figure 21). In addition, MSATs were asked to discuss if the program had any influence on their career aspirations in academic medicine. A total of 17 MSATs (61% of the sample) provided responses to this item. Themes included: a commitment to exploring a career in medical education, enjoyment in mentorship and giving back, engaging in content review, developing teaching skill sets, and participating in curriculum building and teaching innovation.

**Figure 21.**
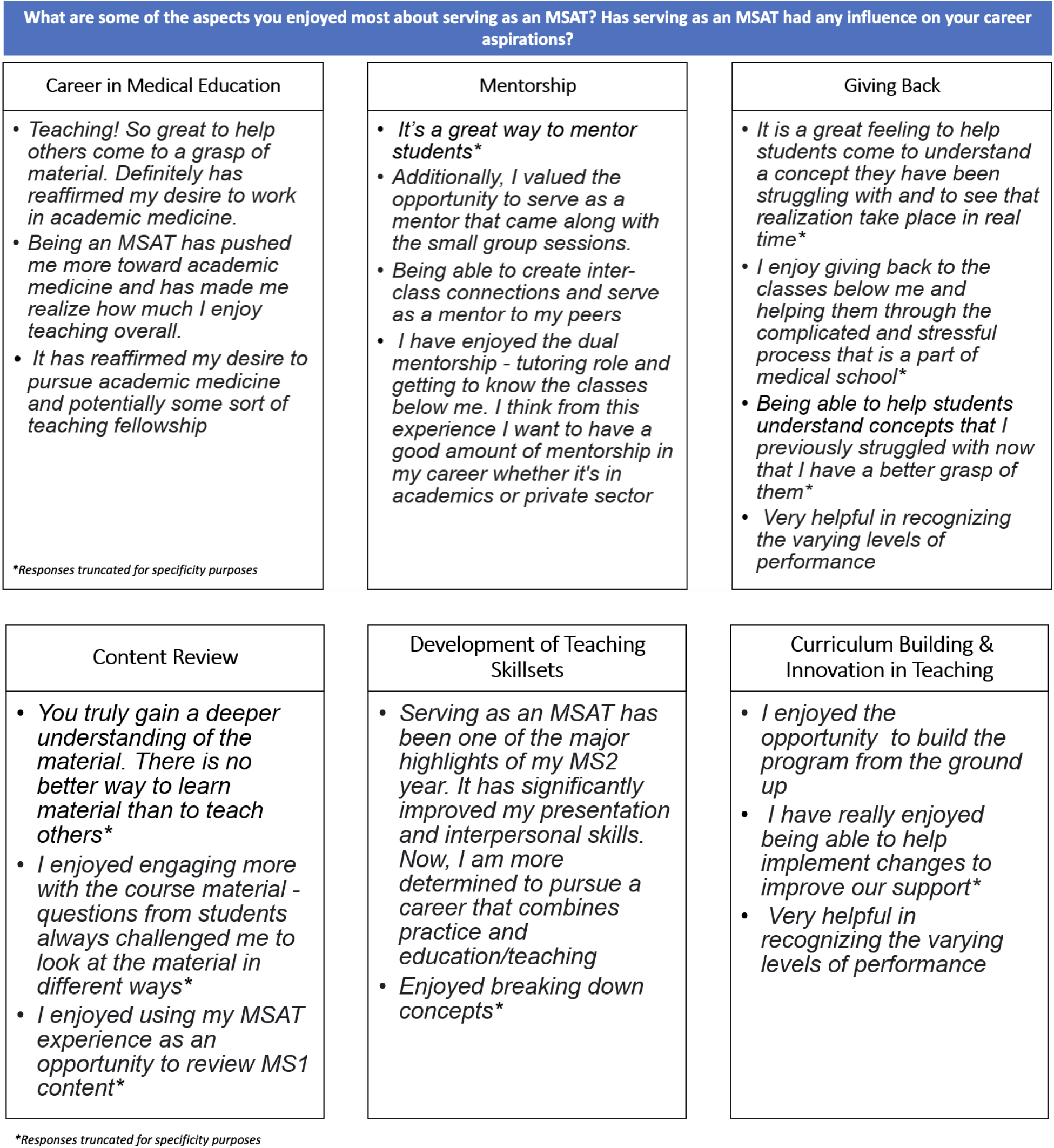
Aspects MSATs enjoyed and influences on career aspirations.

The second open-ended item invited MSATs to discuss ways CLC can improve to better serve them. A total of 15 MSATs (54% of the sample) provided responses to this item. Due to the similarities in the two open-ended items, the third and final open-ended item invited MSATs to share any additional comments or suggestions not captured in the other questions (Figure 22). A total of 3 MSATs (11% of the sample) provided responses to this item. Themes for improvement included: improving teaching materials, providing access to third party resources for MSATs, increasing the number of opportunities for professional networking and development, improving organizational processes and services, and addressing scheduling conflicts.

**Figure 22.**
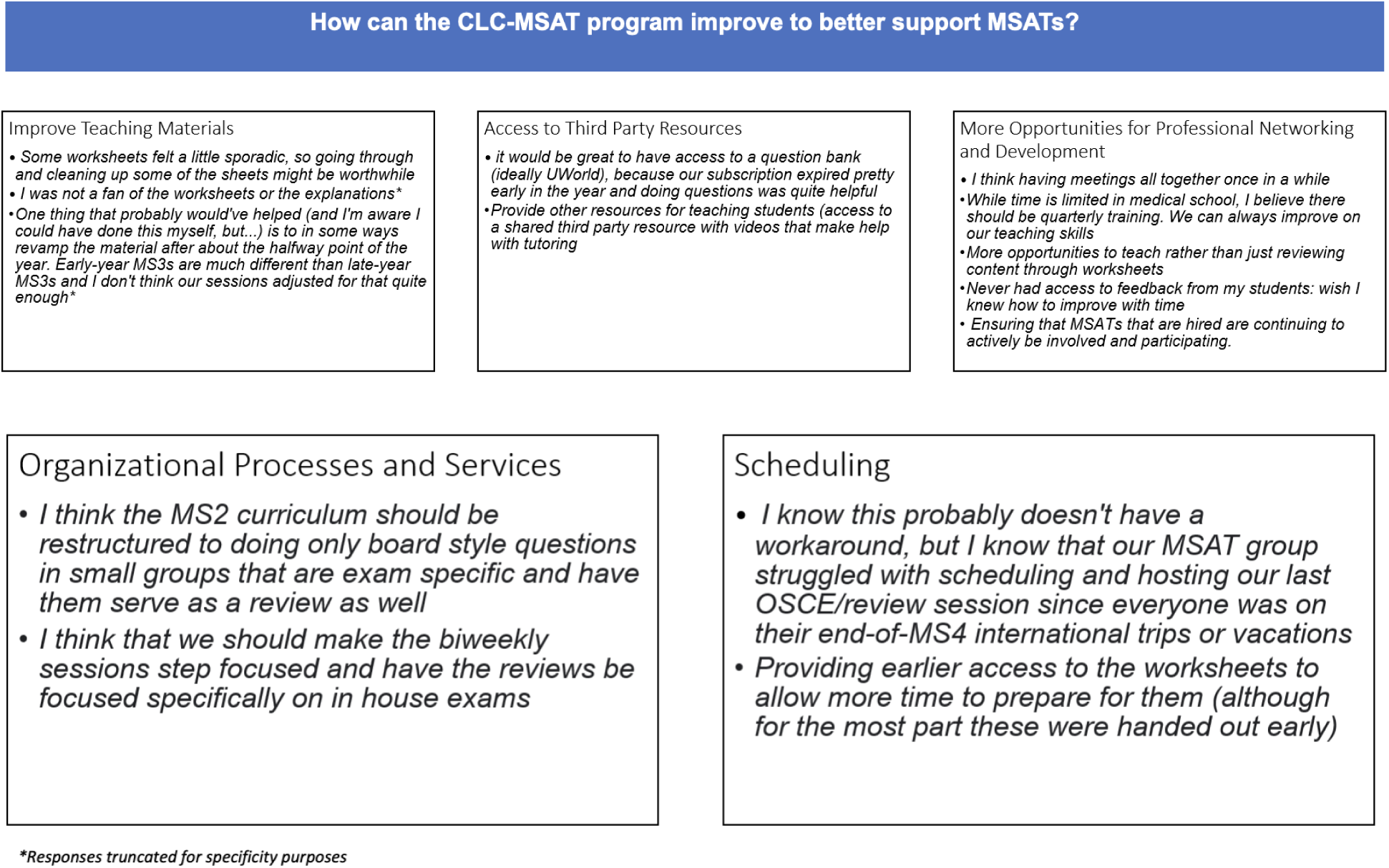
Improvements and general comments from MSATs.

## Discussion

The CLC-MSAT program has been designed to both promote academic excellence for the students in our medical program, as well as to provide authentic teaching experiences for our future physicians. Of particular interest for the researchers was to determine if serving as an MSAT has had any influence on our students’ career aspirations into academic medicine, as well as to determine areas of success and improvement for our program.

Based on our survey results, while 11% of MSATs had no previous teaching experience before CLC, 48% also had no previous teacher training. A total of 100% of MSATs indicated they agreed or strongly agreed they were satisfied with their position, overall. Within these respondents, 96% of MSATs agreed or strongly agreed they would recommend this role to others. An equal number of respondents, 89% each, agreed or strongly agreed that serving as an MSAT has provided them with a stronger grasp of the medical curriculum and made them a stronger teacher. As a theme of teaching efficacy, 100% of MSATs agreed or strongly agreed they worked well with their students.

Our interest was to understand if serving as an MSAT had any influence on a career in academic medicine. These findings indicated 89% of MSATs agreed or strongly agreed they envisioned this career before serving as an MSAT compared to 96% who agreed or strongly agreed after serving as an MSAT. Interestingly, one MSAT indicated they strongly disagreed that they envisioned a career in academic-medicine before CLC; this statistic dropped to 0% of MSATs disagreeing or strongly disagreeing they wanted to be in academic-medicine after serving in CLC.

As we focus on the experiences of our MSATs within the CLC program, it is worthwhile to examine the unique characteristics of our respondents. For example, the 2022-2023 MS1 program was staffed by 18 MSATs, the MS2 program had 14 MSATs, and the MS3 program had 15 MSATs. Within our particular sample, 82% of respondents served in either the MS2 or MS3 programs despite accounting for only 62% of the overall number of MSATs in the program. Thus, the majority of our responses were derived from respondents in the clerkship phase of their medical school journey.

This is an important distinction to highlight, as students in the latter phase of their medical school journey may have more concrete career plans than students in their pre-clerkship years. As such, these upper class-standing students may have joined the CLC program with clearer intentions as to how this program could facilitate their career aspirations compared to pre-clerkship MSATs who may still be in an exploratory phase of their post-graduation plans.

Additionally, 61% of respondents indicated they held a student leadership role in CLC, which would constitute a director or specialist title. A total of 18 students, or 40% of MSATs, held a leadership title for this iteration of CLC. The high representation of student leaders in these responses may have skewed the overall representation of MSATs’ experiences. Essentially, the MSATs who sought and executed leadership roles tended to be students who vocalized their desire to pursue academic medicine after CLC, which was a strong factor as to why their application was originally accepted for the position.

In lieu of these considerations, we are pleased to see a high overall satisfaction rate for our MSATs in addition to sentiments that serving in this program has refined or confirmed many of their aspirations to pursue a career in academic medicine.

### Strengths & Positive Outcomes

**Figure 21.**
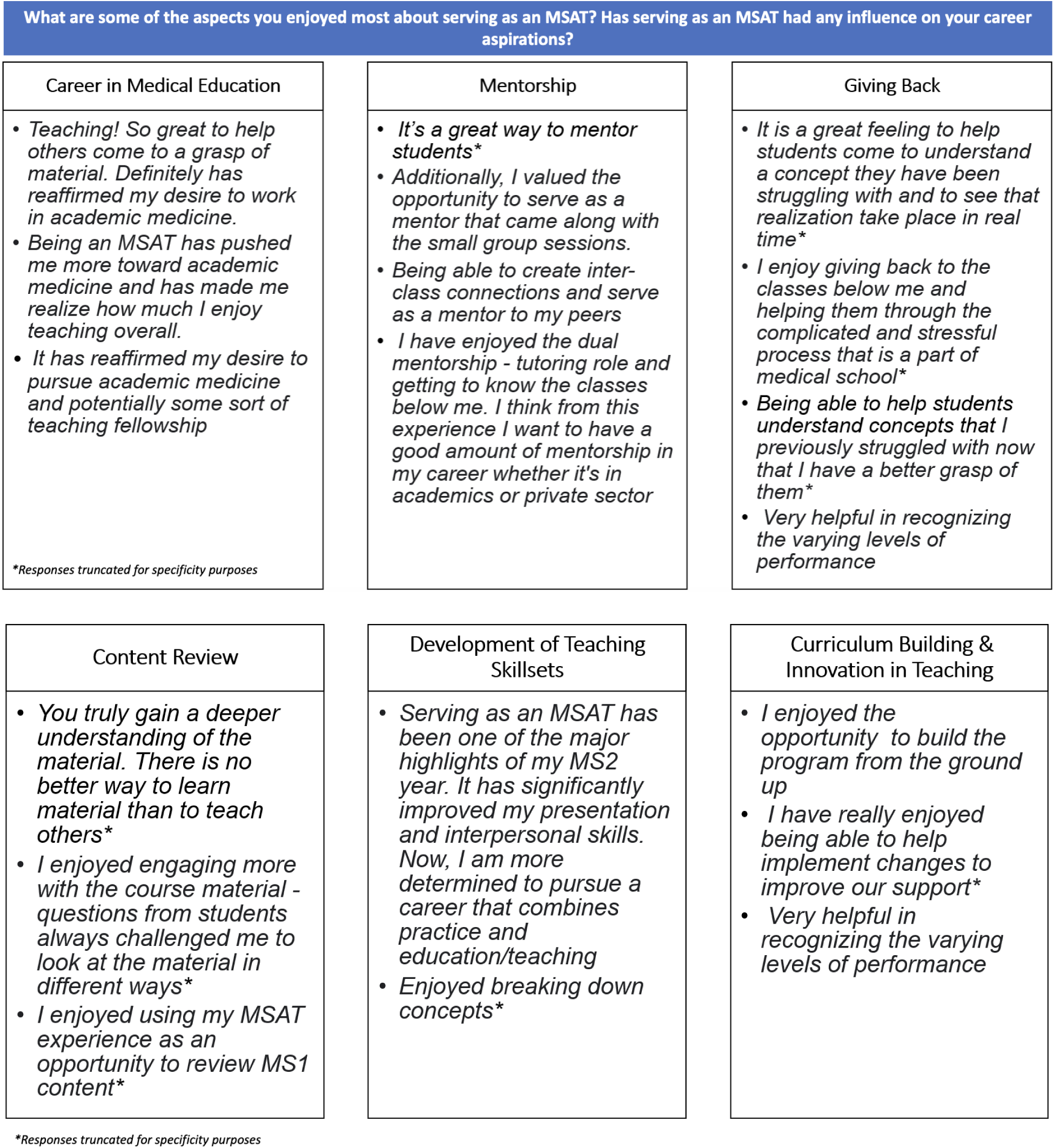
Aspects MSATs enjoyed and influences on career aspirations.

### Career in Medical Education

Before serving in the CLC program, most MSATS (89%) strongly agreed (61%) or agreed (29%) they envisioned a career in academic medicine. However, after serving as an MSAT, 96% of the team either strongly agreed (63%) or agreed (33%) they envisioned a career in academic medicine. Part of the modest 7% increase could potentially be explained by the naturally self-selecting bias of our sample; most MSATs held interests in academic medicine even before CLC employment, which likely attracted them to join the program in the initial phase.

Open-ended responses within the impact CLC has had on career aspirations included the enjoyment MSATs found in helping students grasp difficult content, which reaffirmed their desire to explore academic medicine. The personal, perceived teaching efficacy of helping students master content they once struggled with may have potentially contributed to MSATs’ perspective of being an effective teacher.

### Mentorship

The primary currency for the successful functioning of medicine and medical education is a healthy mentor-mentee relationship. The significance of mentorship in education has been emphasized at multiple levels. It was encouraging to see that MSATs also perceived this as a great opportunity to mentor the next generation.

### Giving Back

Related to teaching efficacy and mentorship attributes included the theme of giving back. Specifically, MSATs developed strong positive associations with assisting struggling students and seeing their development and confidence transform in real-time. MSATs also observed and learned how to recognize students at varying levels of performance to help foster teaching pedagogy to meet the students where they were currently at.

### Content Review

A natural benefit to the tutor-tutee relationship is for the tutor to gain a deeper understanding of challenging material in the effort to teach it to another student. MSATs reported using their position as an opportunity to review previous material and to master the content in a way they had not been able to previously as a student. Additionally, student questions allowed for MSATs to view material under different lenses, further strengthening their grasp of medical content.

### Development of Teaching Skill Sets

CLC was reported to be the highlight of one of the MSAT’s second year of medical school. Specifically, one respondent indicated the program helped improve their presentation and interpersonal skills. Combined, these factors motivated the MSAT to pursue a career that combines medical practice with education and teaching.

### Curriculum Building & Innovation in Teaching

The opportunities for innovating and improving the CLC program were reported as highlights for several MSATs. For example, MSATs discussed how they were able to help build the program “from the ground up” and participated in implementing changes to improve service offerings. MSATs take an active role in executing and improving CLC, which was meaningful for them to recognize.

## Feedback and Improvements

As evidenced by our ongoing surveys, our program is invested in supporting our MSATs by using their feedback to inform our services. Each of these items are outlined below based on the qualitative data captured on our survey.

### Improve Teaching Materials

Each year of CLC builds upon materials from the previous year. Our materials consist of worksheets with board style questions that are derived from influences of existing resources (practice question banks) and in-house vignettes co-designed by our faculty. A new process addition to help improve our teaching materials is for the student curriculum specialists to send their materials to the staff director of CLC, as well as to the faculty course director of the particular content area. While the faculty course director assesses content accuracy and high yield appropriateness, the staff director makes recommendations on learning objectives, organization, and scaffolding opportunities to meet diverse learning levels of students. Additionally, most CLC materials pass through a review of at least five MSATs for proofreading.

### Provide Access to Third Party Resources

In 2023, UCISOM provided free vouchers to the practice question bank, UWorld, to all second year medical students. Due to the costs associated with this offering, it is currently unclear if the university will be able to provide the same access to third party resources for all levels of medical students. This is an area of consideration which will be revisited in the new fiscal year.

### Increase Opportunities for Professional Networking and Development

In response to requests for more opportunities for professional networking and development, the staff director of CLC-MSAT instituted the following changes and mandatory trainings for all MSATs: introduced diversity, equity, and inclusion (DEI) sessions, mental health workshops, as well as improved initial and ongoing teaching pedagogy instruction.

Two DEI trainings are hosted by DEI experts within the UCISOM and UCI main campus departments. The sessions focus on implicit bias, cultural competency, identity salience, and intersecting identities as they relate to strategies and techniques for effective cross-cultural mentoring and advising. Mental health workshops are hosted by the UCI Counseling Center, which includes a suicidal ideation certification program for our MSATs, referred to as Question, Persuade, Refer (QPR), to identify students at risk for mental health challenges. The initial and robust teaching training took place before the start of the 2023-2024 CLC-MSAT program, which was hosted by the staff director of CLC, who is also an adjunct STEM professor and received teaching credential training for secondary education. Additional pedagogy training is hosted in conjunction with UCI’s Division for Teaching and Educational Innovation department to help MSATs adopt and refine their skills for common teaching challenges, such as increasing student participation.

In addition, the number of staff meetings have increased for the 2023-2024 CLC-MSAT program in which weekly, biweekly, and monthly meetings are held with specific MSATs on the team. For example, weekly meetings are held between the student program directors and the staff director of CLC, biweekly for MSATs in specialist roles, and monthly for all other MSATs.

### Improve Organizational Processes and Services

To address areas of improvement in organizational processes and services, our CLC team has been increasingly explicit in communication regarding CLC positions, time commitments, and expectations. Although we have operated using position descriptions from the pilot year of our program, we have made previously ambiguous information more concrete. For example, we have expanded language from, “the MS1 education research specialist will conduct surveys throughout the year,” to “the MS1 education research specialist will be responsible for revising, distributing, analyzing, and reporting data for the MS1 program every October, December, and February of the academic year.”

In addition, CLC leadership has been more intentional about conveying expectations by role, as individual tutoring sessions have notoriously been the most difficult service to staff. As such, MSATs must confirm their acceptance that they will be expected to serve as a 1:1 tutor in addition to their large group teaching sessions when formally accepting their employment offer. This one change has led to simpler staffing for our individual tutoring sessions compared to previous years.

Another improvement area of organizational processes relates to navigating interpersonal differences between MSATs. For example, although an MSAT may share a leadership title of director or specialist, they still remain in a horizontal reporting structure to other MSATs. To help our team navigate challenges they may inevitably face with one another, we have increased the frequency and transparency of our meetings, as our quality communication channels have helped mitigate previous misunderstandings. The CLC-MSAT team remains a united front, committed to providing robust and inclusive tutoring resources to those we are privileged to serve.

## Future Directions

Our CLC-MSAT program has been synonymous with seeking feedback and adapting to change with timely and relevant support for our MSAT team. While we have introduced new and robust trainings for our team, we are also continuously creating new leadership positions for our MSATs. Our intention is to help our MSATs gain valuable experience in a setting which supports and competitively compensates them. For example, we have created director and specialist-level roles based on MSAT requests, including finance, social media, and board exam responsibilities. Our team regularly conducts research and presents and publishes on our findings to provide ample opportunities for MSATs to gain this invaluable experience.

Future directions for CLC-MSAT include longitudinal research to track where our graduated MSATs find their careers after residency. We are excited to likely be able to determine this data for several of our MSATs from our pilot program in the next two years. Additional areas for CLC include increased marketing of our services to undergraduate students interested in UCISOM to showcase the variety of careers in medicine and the doors that may open as a result of serving as an MSAT.

In order to provide additional pedagogy training to MSATs, we plan to sponsor interested MSATs to attend the annual conference on Educating Learners through Innovation and Technology (ELITe) organized by the Medical Education unit of University of California Irvine School of Medicine. During this two-day conference, the instructors teach principles of effective learning through innovations in instructional technology, curriculum development and design, evidence-based slide design, writing effective learning objectives, gamification of medical education, building DEI and cultural competency into curricula, formative and summative evaluations and program evaluations. Historically, this conference has been attended by residents, fellows and other physicians interested in medical education. Recently, many of the basic science faculty who are teaching pre-clerkship curriculum are also taking advantage of this training. Our plan is to open fee waivers for interested MSATs who would like to further advance and refine their teaching in medical education skills. We hope that this training will help fill the current gap of medical educators in the field.

## Conclusion

As evidenced by CLC’s continued expansion from a team of 18 MSATs in 2020 to 73 MSATs three years later, our attention is equally invested in supporting academic excellence for our students in addition to cultivating skilled, future academic-clinicians. We continuously seek out feedback from our MSATs to support their career trajectories. Results from this feedback include providing new service offerings for our staff, such as DEI, mental health certifications, and initial and ongoing teaching pedagogy. We are responsive to our MSAT needs and committed to providing any support which may help contribute to our MSATs pursuing careers in academic medicine.

## Supporting information

IRB Approval UCI

## Data Availability

All the data presented or used in this manuscript is publicly available as supplementary data.

