## Supplementary material for "From tutor to future educator: Investigating the role of peer-peer tutoring in shaping careers in medical education": IRB Approval UCI

### #3183 - Medical Students As Teachers & Future Educators: Training Program at School of Medicine, University California Irvine

#### Protocol Information

| Review Type | Status | Approval Date | Continuing Review Date |
|---|---|---|---|
| Exempt | Exempt | May 11, 2023 | -- |

| Expiration Date | Initial Approval Date | Initial Review Type |
|---|---|---|
| -- | May 11, 2023 | Exempt |

#### Feedback

Approval Comment

None

##### Project Details

**Project Title (100 words max):**
Medical Students As Teachers & Future Educators: Training Program at School of Medicine, University California Irvine

**Lead Researcher/Investigator:**
Lauren Mikki Stokes

**Lead Unit (i.e., Department, Organized Lead Unit, Center or Institute):**
***IR-7476 - MEDICAL EDUCATION (Lead Unit)***

Kuali Research (KR) follows the KFS Organizational Unit Hierarchy.

ATTENTION! For new submissions, Department Chair (DC) or Organized Lead Unit Director (OLUD) sign-off in KRP is required before final committee approval will be granted. For more information, visit the listserv.

##### Submission Screener

**Submission Type:**

IMPORTANT! Be sure to select the correct 'Submission Type'. When 'Submission Type' is changed, the contents of the form will be cleared and replaced with a set of new questions specific to the submission type.
Administrative Determination or Registration

**Submission for Administrative Determination or Registration:**
Exempt Self-Determination

**Lead Researcher's primary school/department/program is:**
Biomedical (Health Sciences)

**Is the project limited to review of records (e.g., educational) and/or biospecimens (e.g., discarded tissues) without subject contact?**
No

##### Project Funding

**Select the funding source(s) (check all that apply):**
No Campus or Extramural funding

##### Exempt Self-Determination

UCI IRB review is required for most activities that constitute engagement in human subjects research, as federally defined.

At UCI, researchers, including undergraduate students proposing exempt human subject research as part of the Undergraduate Research Opportunities Program (UROP) are permitted to self-determination their exempt research without confirmation from the IRB.

The IRB will NOT review this submission. Instead, it will be registered as 'Exempt' research and a confirmation email will be sent.

Should the study sponsor require evidence of IRB review for an exempt self-determination, please provide the sponsor this letter.

- IMPORTANT! All exempt self-determinations must be tracked in Kuali Research Protocols (KRP).
- The Lead Researcher (LR) is responsible for maintaining all documentation related to the Exempt Self-Determination in a separate research record. This documentation may be requested by Human Research Protections (HRP) for quality assurance review. Documentation includes:
  - A printout/PDF of the completed self-determination form.
  - All supplemental documentation, as prompted in the form.
- IGNORE! If prompted to attach documents, please ignore. These prompts are intended for IRB submissions only. Instead, please maintain the documentation on file as noted above.

**Does this study involve the creation, use, or disclosure of Protected Health Information (PHI) (i.e. medical records)?**

List of 18 PHI Identifiers from medical records/clinical encounters:
1. Names
2. Social Security Numbers
3. Dates (including birth date, admission date, discharge
4. date, date of death and exact age if over 89)
5. Medical record numbers
6. Address
7. Health plan numbers
8. Phone numbers
9. Fax numbers
10. Email address
11. Account numbers
12. License/Certificate numbers
13. Vehicle ID numbers
14. Device identifiers/Serial numbers
15. Web URLs
16. IP address numbers
17. Biometric identifiers
18. Facial Photos/Images
19. Any other unique identifier

No

**Does the research involve targeted recruitment of any of the following populations?**

- Children (under 18 years old)
- Adults (age 18 or older) who may not be legally/mentally/cognitively competent to consent
- Prisoners (*may* include parolees)
- American Indian/Alaska Native tribes
- Undocumented people

No

**Are research subjects and/or performance sites located outside the United States? Is this international research?**
No

**Does this research involve a request for UCI to serve as IRB of Record for a non-UCI investigator and/or site engaged in human subjects research?**
No

**Does the Lead Researcher or any study team member have a Disclosable Financial Interest?**

No

IMPORTANT! If all research activities qualify for one or more of the categories below, the study is eligible for exempt self-determination.

**Select the category(ies) that apply to the research:**

Category 1: Education (the following criteria must be met)

Category 2: Research that includes only interactions involving educational tests (cognitive, diagnostic, aptitude, achievement), survey procedures, interview procedures, or observation of public behavior (including visual or auditory recording) if at least one of the following criteria are met:

One of the following criteria must be met:
2i) The information obtained is recorded by the investigator in such a manner that the identity of the human subjects CANNOT readily be ascertained, directly or through identifiers linked to the subjects
2ii) Any disclosure of the human subjects' responses outside the research would NOT reasonably place the subjects at risk of criminal or civil liability or be damaging to the subjects' financial standing, employability, educational advancement, or reputation

Category 3i: Research involving benign *behavioral* interventions in conjunction with the collection of information from an adult subject through verbal or written responses (including data entry) or audiovisual recording if the subject prospectively agrees to the intervention and information collection and at least one of the following criteria is met:

One of the following criteria must be met:

Category 4: Secondary research for which consent is not required: Secondary research uses of identifiable private information or identifiable biospecimens, if the following criterion is met:

###### Self-Determination Eligibility

**The activities, as indicated above, meet the criteria for Exempt Self-Determination.**

**UCI IRB approval is NOT required.**

If your project changes in ways that may affect this determination, please contact the HRP staff for additional guidance.

##### Clinical Trials

###### Clinicaltrials.gov

Registration on ClinicalTrials.gov may be required if one (or more) of the following is true:

- Study is NIH funded and meets the NIH definition of a clinical trial
- Study is DoD funded and registration is required by your specific program
- Study meets the International Committee of Medical Journal Editors (ICMJE) definition of a clinical trial

**Does this research meet the definition of a clinical trial that requires adherence to Clinicaltrials.gov (CT.gov)?**
No

##### Non Technical Summary

**Provide a non-technical summary of the project that can be understood by non-scientists (250 words max):**
A survey will be administered to medical student tutors to determine their satisfaction with their position and to indicate how we can improve our tutoring program.

##### Other UCI Committee Reviews

###### Potentially Hazardous Materials

**Does the research involve human/primate blood, tissue, fluids or primary cells?**
No

##### Study Team

- Lead Researcher (LR): The LR must meet LR Eligibility requirements or have a Faculty Sponsor (FS) listed who is eligible.
- Co-Researcher (CR): CRs are key personnel for conducting the research study. These individuals work closely with the LR to design, conduct, and/or report on the research.
- Research Personnel (RP): Review the RP Heat Map to determine whether they should be listed below.
- Administrative Contact (AC): Add ACs only in the Permissions tab. Do NOT list below.

All study team members must complete the following Collaborative Institutional Training Initiative (CITI) trainings:

- Human Subjects Research Protections and
- Good Clinical Practice, as applicable

Incomplete or expired CITI training will delay IRB approval. For more information, visit HRP Training and Education.

**Researcher:**
Lauren Mikki Stokes

**Email:**


To promote the objectivity of the research, all researchers are required to disclose their related disclosable financial interests, per the IRB COI Policy. If you have any questions about the COI process in general, contact the COI team.

**Each member of the study team for this protocol must be asked the following question to comply:**

**"Do you, your spouse/registered domestic partner, and dependent children have any disclosable financial interests* (i) that would reasonably appear to be affected by this research study; or (ii) in entities whose financial interests would reasonably appear to be affected by this research study?"**
No

Training

| Biomedical Investigators - Basic Course | 01/22/22 - 01/21/27 |
|---|---|

**Degree:**
Other

**Degree Other:**
EDD

**Position/Title:**
Director of Academic Support

***IR-7476 - MEDICAL EDUCATION (Lead Unit)***

**Affiliation:**
UCI Staff

**Researcher Role:**
Lead Researcher

**Permissions:**
Full Access

**Duties:**
Oversight of Research
Finalize Informed Consent
Research Procedures (specify below)

**Specify which research procedures:**
Co-create the survey and analyze data

**Specify relevant training and experience for the referenced duties/responsibilities:**
Researcher and staff

**Researcher:**
Harinder Singh

**Email:**


To promote the objectivity of the research, all researchers are required to disclose their related disclosable financial interests, per the IRB COI Policy. If you have any questions about the COI process in general, contact the COI team.

**Each member of the study team for this protocol must be asked the following question to comply:**

**"Do you, your spouse/registered domestic partner, and dependent children have any disclosable financial interests* (i) that would reasonably appear to be affected by this research study; or (ii) in entities whose financial interests would reasonably appear to be affected by this research study?"**
No

Training

Harinder Singh has no training courses on file.

**Degree:**
PhD

**Position/Title:**
Assistant Adjunct Professor

***IR-7476 - MEDICAL EDUCATION (Lead Unit)***

**Affiliation:**
UCI Faculty

**Researcher Role:**
Co-Researcher

**Permissions:**
Full Access

**Duties:**
Oversight of Research
Access/Analyze Identifiable Information

**Specify relevant training and experience for the referenced duties/responsibilities:**
Researcher and faculty member

##### Background & Purpose of Research

**Describe the purpose, specific aims or objectives and specify the hypotheses or research questions to be studied:**

The purpose is to administer a survey to medical student tutors to determine their satisfaction with their position and to indicate how we can improve our tutoring program. We hypothesize the tutoring program has increased medical students' interest in a career within academic medicine.

##### Subject Population(s)

**Targeted subject populations/data sources (check that apply):**
Adults Competent to Provide Informed Consent
UCI Students/Staff/Faculty

###### Sample Size

**Specify the maximum and expected numbers of individual-level information and/or biospecimens to be accessed/analyzed within each category/group.**

**Will this study take place only at UCI (i.e., does NOT involve other non-UCI sites)?**
Yes

**Sample Size Determination:**

1. **Explain how the overall target sample size was determined** (e.g., power analysis; precision estimation). **Power analysis should (at least) match the primary outcome/endpoint.**
2. **Provide justification of the effect size for the primary outcome based on preliminary data, current knowledge/literature and/or cost consideration.**
3. **If appropriate, provide sample size justification for secondary outcomes.**

70 medical student tutors at UCI

###### Eligibility Criteria

IMPORTANT! If utilizing UCI Health Enterprise Data & Analytics services, include specific timeframes for each eligibility factor, as applicable.

Example:
1. Birth sex: female
2. Age: >= 18 years old as of 2020-01-01
3. The result of the most recent SARS-CoV-2 test (of any type), performed between 2020-01-01 and 2020-12-31, was positive
4. With any sub-classification of type 2 diabetes (E11*) diagnosed at any date prior to 2020-01-01
5. Did NOT have an ED visit between 2020-01-01 and 2020-12-31

**Inclusion/Exclusion Criteria: Identify the factors for limited eligibility and provide a scientific rationale.**

**Is eligibility based on age, gender, pregnancy/childbearing potential, social/ethnic group, or language spoken (e.g., English Speakers only)?**
No, Subject Eligibility is not based on these factors

REQUIRED! This study does NOT include Non-English Speaking Participants (NESP) Revise the above question to 'Yes', and add a scientific justification/rationale for the exclusion of NESP.

##### Pre-Screening for Recruitment

**Will identifiable information be accessed/obtained for the purpose of screening, recruiting, or determining eligibility of prospective subjects?**
No

##### Recruitment Methods

IMPORTANT! Recruitment materials must adhere to UCI Recruitment Guidelines. Various templates are available here: IRB Forms → Recruitment Templates.

**Indicate all methods that will be used to recruit subjects for this study:**

**Recruitment Method:**
Other recruitment methods

**Specify 'Other' recruitment methods:**
Existing medical student tutor staff

##### Informed Consent Process

**Identify the methods of Informed Consent or assent process as applicable for each participant population (check all that apply):**
Oral/Implied informed consent (no signature)

###### Oral/Implied Informed Consent/Assent

**Is this consent method used for all subjects?**
Yes

REQUIRED! Develop and use a Study Information Sheet.

UCI IRB requires ALL of the following:
- An IRB-approved Study Info Sheet will be presented prior to administering research procedures,
- Subjects verify they meet the eligibility criteria,
- Subjects indicate their willingness to participate in the research (e.g., click "Yes")

###### Circumstances of Consent

**Indicate the location where the consent process will take place (check all that apply):**
Internet

**Specify how the research team will assure that subjects, their parents, or their legally authorized representative (LAR) have sufficient time to consider whether to participate in the research:**
Subjects or their LAR will be allowed to take home the unsigned consent form for review prior to signing it

##### Project Locations

**Check all sites where UCI investigator(s) will conduct research activities (e.g., recruitment, informed consent, and research procedures including accessing identifiable, private information about participants):**
Non-UCI Locations (i.e., not UCI, UCI Health, or UCI owned/leased space)
UCI Campus Facilities or Sites (e.g. school, lab, etc.)
UCI Health Facilities or Sites (e.g. hospital, clinics, etc.)
Virtual Locations (e.g. Amazonturk, Zoom, Telehealth/Virtual Care)

**Is the location(s) public or private?**
Public (e.g. coffee house, public park, library)
Private

###### Private Non-UCI Locations

**Specify the location where UCI IRB approved research will occur (check all that apply):**
Other Site(s)

**Specify location(s) or specify group name & provide website link if virtual:**
Remotely at home

**Will personnel at the non-UCI locations engage in research activities?**
No

##### Project Procedures

**Research Procedures (check all that apply):**
Surveys/Questionnaires/Interviews/Focus Group

**Provide a detailed chronological description of the procedures:**

Medical students will be asked to complete a survey about their experience as a tutor. Then, the researchers will analyze the data.

**Specify the total duration of a subject's participation in the study and clearly outline the duration of participation for each study visit and sub-study, as applicable:**

10 - 15 minutes to complete the survey.

**List all data collection tools (e.g., measures, questionnaires, observational tool) below; include citations for standardized/validated measure(s):**

Student satisfaction survey.

**Are all data collection tools standardized or validated?**
No

IMPORTANT! For data collection tools that are not standardized or not validated, maintain a copy of the final version on file in the researcher's record.

##### Risk Assessment

###### Certificate of Confidentiality

**Is the research partially or wholly funded by National Institutes of Health (NIH), including NIH Institutes and Centers?**
No

**Does the research involve a sensitive health-related topic that collects names or other identifiable, sensitive information pertaining to subjects?**
No

###### Potential Benefits

**Is there the prospect of a direct benefit anticipated for subjects?**
No

**Specify the expected potential societal/scientific benefit(s) of this study:**
Providing better services to our student tutors.

##### Alternatives to Participation

**Describe the alternatives to participation in the study available to prospective subjects (check all that apply):**
No alternatives exist. The only alternative to study participation is not to participate in the study

##### Participant Compensation

Compensation is when participants are paid for their time & efforts in research.

- Compensation should be offered on a prorated basis when the research involves multiple sessions.
- For additional information about researcher's/department's responsibilities and current Accounting procedures, see UCI Policy Sec. 701-03.

For more information see: Compensation Info.

**Are participants compensated?**
No

##### Confidentiality of Research Data

###### Participant Identifiers

**Will subject/patient identifiers be collected or retained?**
No

###### Info/Biospecimen Storage

**Indicate how information and/or biospecimens (including signed consent forms) will be stored (check all that apply):**
Information will be maintained electronically. Information will be password protected and maintained in an encrypted format

Encrypted Format

**Specify where the information will be maintained electronically:**
Google Forms

Info/Biospecimen Retention

**Indicate how long research information/biospecimens will be retained:**
This research includes the potential for future secondary research using information/biospecimens which will be stored and maintained indefinitely

Info/Biospecimen Sharing

**The research team, authorized UCI personnel, the study sponsor (as applicable) and regulatory entities such as the Food and Drug Administration (FDA) and the Office of Human Research Protections (OHRP), may have access to participants' study records to protect their safety and welfare.**

Sharing Within Scope of Project

**Will research materials (information/ biospecimens) be shared with collaborators (i.e., researchers not covered under the UCI project), for purposes within the scope of the current project?**
No

Sharing Outside Scope of Project

**Will information and/or biospecimens be shared, used again, or stored for undefined future research purposes beyond the scope of the current protocol?**
No

Need Help?

**09/27/2022 UPDATE: There is a slight time delay (30+ seconds) when "submitting" a transaction. Please do not refresh or close the page. The transaction will eventually go through.**

**Kuali is currently working to resolve the performance issues our customers are experiencing. Thank you for your patience and your partnership.**

**Contact the Office of Research**

**For KRP technical questions or issues:**
- Visit the KRP User Guide
- Contact Electronic Research Administration (ERA)

**For IRB questions and regulatory or institutional guidance:**
- Visit Human Research Protections (HRP)
- Contact the HRP staff

#### Administrative Details Form

Project Status

**Committee:**

**Project Status:**

**Date of Action/Determination:**

**Amendment Status:**

**Date of Amendment Action/Determination:**

**ERA Transcription Date:**

*Not applicable for new studies submitted after September 07, 2021*

**Pre-2018 Common Rule:**

*Not applicable for studies initially approved after January 21, 2019*

**Date of Transition:**

*Not applicable for studies initially approved after January 21, 2019*
